# Task-Oriented Predictive (Top)-BERT: Novel Approach for Predicting Diabetic Complications Using a Single-Center EHR Data

**DOI:** 10.1101/2024.04.15.24305843

**Authors:** Humayera Islam, Gillian Bartlett, Robert Pierce, Praveen Rao, Lemuel R. Waitman, Xing Song

## Abstract

In this study, we assess the capacity of the BERT (Bidirectional Encoder Representations from Transformers) framework to predict a 12-month risk for major diabetic complications—retinopathy, nephropathy, neuropathy, and major adverse cardiovascular events (MACE) using a single-center EHR dataset. We introduce a task-oriented predictive (Top)-BERT architecture, which is a unique end-to-end training and evaluation framework utilizing sequential input structure, embedding layer, and encoder stacks inherent to BERT. This enhanced architecture trains and evaluates the model across multiple learning tasks simultaneously, enhancing the model’s ability to learn from a limited amount of data. Our findings demonstrate that this approach can outperform both traditional pretraining-finetuning BERT models and conventional machine learning methods, offering a promising tool for early identification of patients at risk of diabetes-related complications. We also investigate how different temporal embedding strategies affect the model’s predictive capabilities, with simpler designs yielding better performance. The use of Integrated Gradients (IG) augments the explainability of our predictive models, yielding feature attributions that substantiate the clinical significance of this study. Finally, this study also highlights the essential role of proactive symptom assessment and the management of comorbid conditions in preventing the advancement of complications in patients with diabetes.

## Introduction

Micro and macro-vascular complications induced by diabetes can have substantial impact on diabetes management and patient care^1,2^. Early prediction of these complications allows for the identification of high-risk patients and active implementation of preventive measures^3–7^. With this motivation, researchers have developed models predicting diabetes-related complications, primarily emphasizing cardiovascular outcomes and, to a lesser extent, kidney and eye complications^3,8^. However, most of the prior research was focused on predicting risk scores using a limited number of risk factors, often curated from previous literature ^8–14^. Despite many machine learning (ML) and deep learning (DL) models that emerged in recent research, classical ML models dominated these studies– mostly limited to performance comparisons, with only a minority delving into exploring novel risk factors and discovering new knowledge^15,16^.

Digital patient data from electronic health records (EHR) systems play a crucial role in developing clinical risk prediction models, thereby guiding the development of robust, evidence-based medical interventions^13,17,18^. Structured EHR systems systematically documents the timeline of patient encounters, encompassing elements such as demographics, vital signs, diagnoses, prescribed medications, lab test results, and medical procedures. Hence, feature vectors derived from EHR data can enable the use of traditional ML and DL techniques^19,20^. However, the intricate and abundant information contained within EHR data is often condensed to create summary features for predictive models. This process can diminish the temporal and contextual richness of the data. This simplification frequently neglects the complex nature of EHR data, such as sparsity, heterogeneity, and irregular patterns of patient visits, leading to model overfitting and lack of model generalizability^21^.

The resemblance between EHR sequences (time series sequences from different data modalities) and natural language (word sequences) has led to the adoption of advanced NLP (natural language processing) techniques for EHR data. Convolutional neural networks (CNN)^22^ and recurrent neural networks (RNN)^23^ with embedding layers have enhanced the capture of sequential data but have fallen short in recognizing long-term dependencies, hindered by problems like gradient instability. Subsequent techniques, such as Long Short-Term Memory (LSTM) models, have been designed to forecast clinical events, yet they have been hampered by slow training processes and persistent data complexities^24,25^. A transformative breakthrough that revolutionized the learning of contextual and temporal information in language models is the Transformers (2017)^26^ architecture. The pioneering studies^21,27,28^ in applying Transformers to structured EHR data, notably BERT (Bidirectional Encoder Representations from Transformers, 2019^29^), have shown their effectiveness in capturing the complex temporal patterns and navigating the intricacies of EHR for clinical predictions. For instance, BERT’s advanced embedding framework captures the nuanced semantics and context of patient timelines, addressing EHR data sparsity and varying time intervals between encounters. Unlike traditional ML models, BERT transforms these sequences into dense embeddings, preventing learning from sparse matrices of numerous zeros. Moreover, its self-attention and feed-forward mechanisms efficiently learn long-term dependencies and uncover complex event relationships, improving model transparency.

Earlier studies adopted a two-stage process where pretraining on extensive multi-site EHR databases was followed by finetuning on a smaller, specific cohort for clinical predictions. For example, Med-BERT (2021)^27^ was pretrained on a multi-site dataset encompassing 28 million patient records for one week before finetuning on three smaller, distinct datasets. However, patient privacy laws and proprietary data rights present significant obstacles to data sharing^30,31^, thus impeding the distribution of EHR-based pretrained models, deviating from a common practice in the NLP field. This leads us to the question: Could BERT’s unique ability to manage EHR-specific complexities still provide valuable and interpretable predictions when applied to data from a single medical center?

Therefore, in this study, we investigated BERT’s potential in predicting a 12-month risk of developing significant complications, including retinopathy (RET), chronic kidney disease (CKD), neuropathy (NEUR), and major adverse cardiovascular events (MACE) in diabetes patients from a single-center EHR data. Shifting from the conventional pretraining-finetuning paradigm, we introduced an end-to-end training and evaluation method called task-oriented-predictive (Top)-BERT. This innovative approach concurrently optimizes the model for multiple specific prediction tasks––enhancing its effectiveness particularly in settings constrained by limited data.

In Top-BERT, we utilized the sequential input structure, embedding layer, and encoder stacks inherent to BERT to train and evaluate three tasks simultaneously: the conventional Masked Language Model (MLM), a binary classification for prolonged hospital stay (1 if the length of stay >7, else 0), and a multilabel sequence classification for the four complications mentioned above. We aggregated the loss of the three tasks, which was backpropagated throughout the entire network, leading to improved learning of our model in a limited cohort sample size. We evaluated our Top-BERT model against conventional pretraining-finetuning experiments with sequential input. We also compared its performance with traditional ML techniques, such as XGBoost, using a one-hot encoded representation of the features. Our Top-BERT model demonstrated a more effective ability to distinguish between classes and maintained robustness in managing the class imbalance for multilabel outputs, outperforming both the traditional approaches of pretraining-finetuning and XGBoost models.

We also investigated into the embedding structure of BERT, which typically uses positional and segment embeddings, to discern the sequential order of patient histories. Prior studies implemented unique embeddings, such as Med-BERT’s incorporation of visit numbers^27^ and BEHRT’s addition of age to positional and segment embeddings^21^. Our study evaluated the impact of integrating temporal factors as embeddings—such as age, visit sequence, and inter-visit intervals—to predict diabetes complications. We utilized AUROC (area under the receiver operating characteristics curve) and Shannon’s entropy ^32^ for performance comparison, aiming to determine the most informative temporal representation in patient histories for our single-center dataset.

To the best of our knowledge, our study represents a novel effort to apply a modified BERT architecture for predicting four significant micro and macro-vascular complications in diabetes patients within a single research framework while navigating the complexities inherent in EHR data. Furthermore, in our study, we systematically evaluate feature importance across patient visits, identify highly contributing features for each complication, and examine age-specific feature influence variations using Integrated Gradients (IG), a gradient-based feature attribution method optimized for deep learning^33^. Unlike attention mechanisms in models like BERT, which offer partial insights, IG provides a comprehensive analysis by tracing the gradient flow from a predefined baseline to the input. This approach ensures adherence to the axioms of sensitivity and implementation invariance, which are not satisfied by other backpropagation methods such as layer-wise relevance propagation (LRP)^34^ or Deconvolutional networks (DeConvNets)^35^. This methodological adaptation significantly enhances the explainability of our study’s findings within the clinical setting, offering nuanced insights into the factors driving the model’s predictions.

The remainder of this paper is structured as follows. The method section outlines our approach to model derivation and development, data preparation, followed by the detailed experimental setup for this study. The result section presents the comprehensive analysis of the study cohort, model comparison, and analysis of the model explanations. Finally, the discussion section elucidates the methodological advances and clinical implications of our findings and provides a concluding summary at the end.

## Methods

### Model Derivation & Development

#### Conventional BERT framework

The architecture of BERT^29^, initially designed for language representation, is built upon a multi-layer bidirectional Transformer encoder based on Vaswani (2017)^36^. The fundamental elements of BERT encompass (i) input/output representation, (ii) the configuration of embedding layers, and (iii) the architecture of the Transformer encoder layers. In essence, BERT necessitates input sequencing, which involves tokenization using a designated vocabulary and incorporating special tokens like [CLS] at the beginning of the sequence and [SEP] to denote sequence separation. After tokenization, the input token traverses through the embedding layers, each capturing distinct contextual facets of the sequence. This process yields a summed embedding comprising token, segment, and positional embeddings. The embedded input then passes through the Transformer Encoder stack, processing every token simultaneously.

BERT training entails two steps: (i) pretraining tasks and (ii) finetuning tasks. Pretraining involves two self-supervised tasks. The Masked Language Model (MLM) randomly masks a fraction of input tokens and subsequently predicts these masked tokens through a training head atop the encoder stack. Concurrently, Next Sentence Prediction (NSP), a binary classification task, further trains BERT to comprehend inter-sequence relationships. During pretraining, contextualized embeddings are generated for each input token. In the finetuning phase, the pretraining weights are loaded to train the finetuning cohort for specific downstream prediction tasks like classification with an additional prediction head (classification layer) integrated over the encoder stack.

#### Motivation for the input representation for EHR data in BERT

The patient timeline in an EHR is a series of visits, each documented with various health-related elements such as diagnoses, medications, and lab results. Reflecting this, Li et al. (2020)^21^ designed BEHRT, inspired by BERT, using patient diagnoses information per visit to predict future diagnoses, denoting the EHR timeline of each patient *p* with *n*_*p*_ number of visits as, 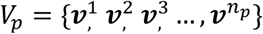, where 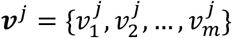 contains ordered clinical entities in the *j*th visit. Similar to the BERT model, they introduced the start of medical history (i.e., [CLS]) and the space between visits (i.e., [SEP]), which results in a new sequence, 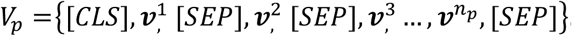. However, Med-BERT(2021)^27^ did not use the specific tokens [CLS] and [SEP] at the input layer due to differences in EHR and text input formats. In BERT, [SEP] serves as a separator between two adjacent sentences for the next sentence prediction task, and as reasoned for Med-BERT, visit embeddings effectively separate each visit in EHR, making the addition of [SEP] redundant. Similarly, Rao-BEHRT (2022)^28^ modified the BEHRT representation of EHR sequences and dropped the [CLS] token. In all these studies, the visit sequence for each patient, which consists of diagnoses/medications, is converted to sequence to represent the temporal structure of EHR.

Moreover, adaptations of the embedding layer for EHR temporality representation varied in the previous studies. The embedding layer in BEHRT^21^ incorporates four types of embeddings: disease, position, age, and visit segment. Positional encodings enable the network to capture positional interactions among diseases using a pre-determined encoding addressing the imbalanced distribution of sequence length in EHR. Age serves as a risk factor for diseases and provides chronological information, while visit segment indicates the separation between visits and differentiates adjacent visits of a patient. Similarly, Med-BERT (2021)^27^ utilized diagnosis code embeddings, visit embeddings, and serialization embeddings to capture clinical code representations, distinguish visits, and capture code order. Additionally, Rao-BEHRT (2022)^28^ used encounter (disease/medication), age, and calendar year. Moreover, Med-BERT was trained on structured diagnosis data using ICD codes, unlike BEHRT and Rao-BEHRT (2022)^28^, which used Caliber codes (developed by a college in London).

#### Our proposed Top-BERT architecture

Our designed Top-BERT, leveraging the foundational components of BERT, serves as a versatile end-to-end training and evaluation architecture that can be tailored directly for a wide range of clinical predictive tasks. Top-BERT utilizes input representation, embedding layer, and encoder stacks like traditional BERT. The input sequence for Top-BERT represents the EHR sequence for each patient *p* with *n*_*p*_ number of visits as: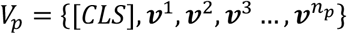, where 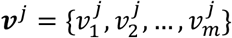 contains the ordered clinical entities in the *j*th visit. We added the [CLS] token at the start of every sequence, which is essential for training BERT for classification tasks. During training, the final hidden state of the [CLS] token becomes the summary representation of the sequence. Due to BERT’s self-attention layers, the [CLS] token integrates context from the full sequence, resulting in a complete summary by the last layer, making it suitable for sequence-level classification tasks. In Top-BERT’s embedding architecture, we examined different temporal factors such as age, number of visits, time between visits, and the conventional positional and segment embeddings to best represent patient timelines in EHRs. The specifics of these experiments will be elaborated in our experimental design section.

Figure 1 shows the modified BERT architecture used for training Top-BERT using the conventional terminology of the model. In the standard BERT pretraining^29^, the input embeddings for *i*th token in *j*th visit, 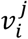 represented as ***e***_*i*_ ∈ ℝ^*hidden*_*size*^, passing through the *BERT* model, get transformed into context vectors and then into hidden states by the BertAttention layer. These are further processed by BertIntermediate’s dense layer and normalized by BertOutput. The BertEncoder stack, comprising multiple BertLayers, yields encoded outputs for all encoder layers (*all_encoded_layers*). The BertModel produces two primary outputs: *all_encoded_layers* and *pooled_output*, the latter derived from the BertPooler function applied to the hidden state of the initial [CLS] token. The *pooled_output* is essential for training the model for any classification tasks by adding an appropriate dense layer to output logits.

**Figure 1A.**
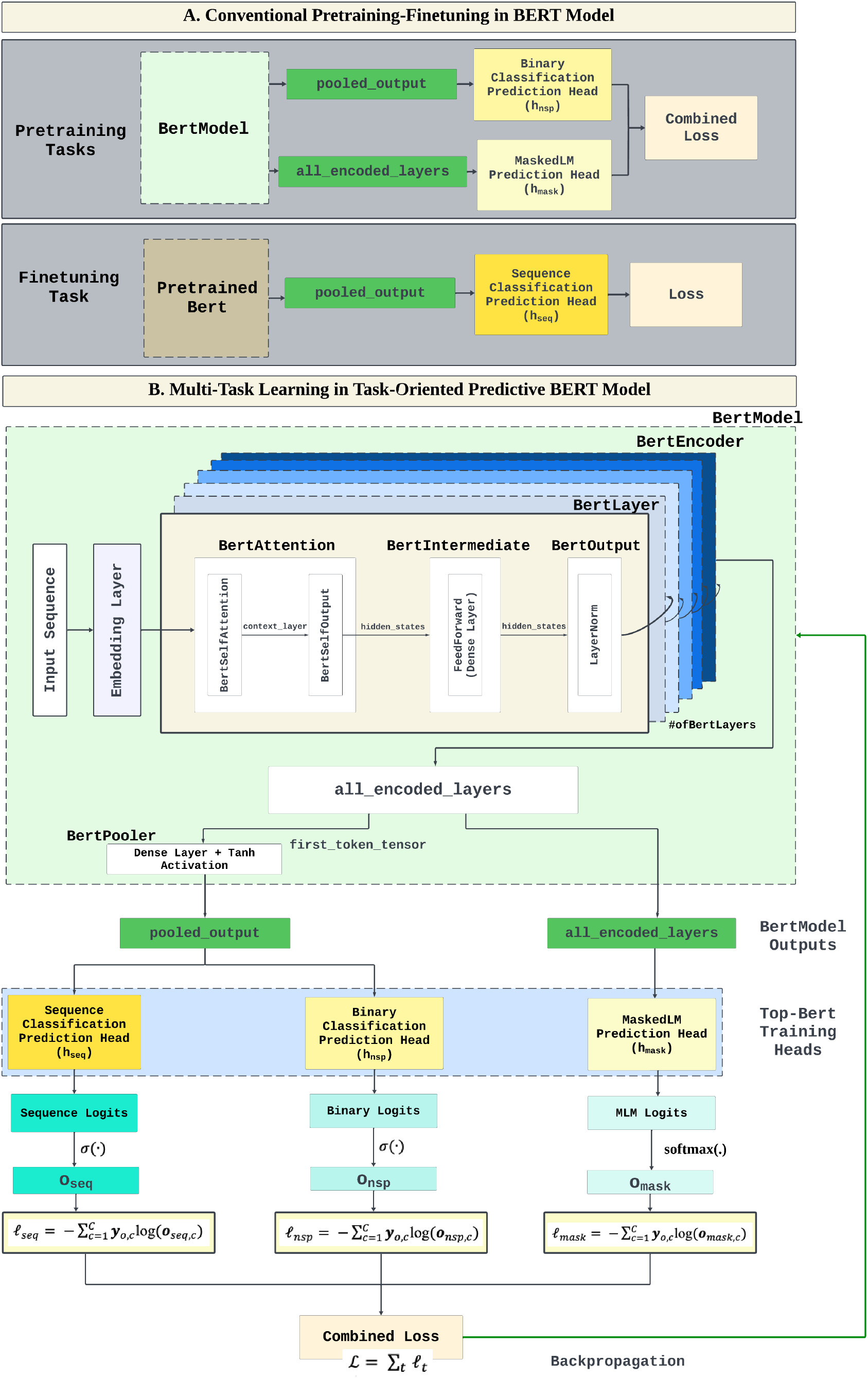
This figure shows the conventional pretraining-finetuning steps in BERT framework. During pretraining, BERT employs two self-supervised tasks: Masked Language Model (MLM) and Next Sentence Prediction (NSP), which discerns relationships between sentence pairs. For finetuning, these pretrained weights initialize a model tailored to specific tasks. Figure 1B. An infographic showing the task-oriented predictive BERT framework using the conventional terminology in BERT models. In BERT’s architecture, embedded inputs pass through BertAttention layers to generate context-sensitive hidden states, further refined by a series of BertLayers to produce a stack of encoded outputs. The model outputs both these encoded layers and a pooled_output–the latter obtained from the hidden state of [CLS] token. We enhanced the BERT architecture and blended pretraining and finetuning into a unified process by integrating a sequence classification head. Utilizing pooled_output from the BertPooler, our adapted model conducts end-to-end training for sequence classification tasks. This method leverages multitasking during pretraining, allowing for simultaneous loss optimization across tasks and enhancing the model’s predictive performance.

In the MLM task, for a masked token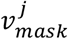, the model output before the activation layer can be represented as:

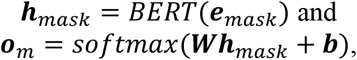

where ***e***_*mask*_ is the embedding of the masked input,***h***_*mask*_ is the output of *BERT* model for the masked token, ***W*** is the weight matrix of the output layer, ***b*** is the bias term, and ***o***_*m*_ is the predicted probabilities.

Similarly, for the NSP task, for a token 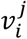, the model output is represented as:

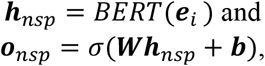

where ***e***_*i*_ is the embedding of the input, ***h***_*nsp*_ is the hidden state of [CLS] token (*pooled_output)*, ***W*** and ***b*** are the weights and biases for NSP, and ***o***_*nsp*_ is the predicted probabilities. *σ*(·) is the sigmoid activation function.

As a novelty of our approach, we have augmented the BERT architecture with a sequence classification head, merging the pretraining and finetuning steps into one end-to-end training and evaluation process. While the original pretraining heads focus on BERT’s standard tasks, the new head utilizes the *pooled_output* to yield precise logits for sequence classification (dense layer to output logits). For this sequence classification task, we define the output as:

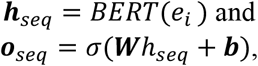

where ***h***_*seq*_ is the hidden state of [CLS] token (*pooled_output)* and *o*_*seq*_ is the predicted probabilities. Thus, Top-BERT can be represented as:

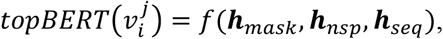

where *f*(·) represents the multitask model with self-supervised (***h***_*mask*_) and semi-supervised (***h***_*nsp*_), and supervised (***h***_*seq*_) tasks.

This enhancement aligns with BERT’s original design and extends its utility by introducing multitasking capabilities within the pretraining phase itself. Each task’s logits undergo their respective loss functions, and the cumulative loss from the three tasks is backpropagated, updating the network’s weights. The combined loss can be represented as:

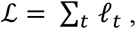

where *t* ∈ {*MASK, NSP, SEQ*} and each 𝓁_*t*_ can be represented a s a summation o f each task-oriented cross-entropy loss:

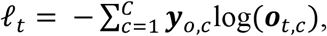

where *C* is the number of classes, *y*_*o*_ is the one-hot encoded true label, and *o*_*t,c*_ is the predicted probability for class *c*.

This results in training and evaluating the BERT model for specific prediction tasks, refining its capability by simultaneously optimizing it for multiple objectives. This multitask optimization could be pivotal in domains with limited data or where task-specific nuances are critical, offering a more nuanced and direct path to task-specific model refinement. We will discuss the implementation of Top-BERT for diabetes-related complication prediction tasks in the following subsections.

### Data Preparation

#### Data Source

Our research utilized the EHR database from the University of Missouri (MU) Hospital, which contains over 1.25 million patient records reflecting a wide geographic and demographic diversity range. MU follows the PCORnet Common Data Model (CDM) for structuring and representing their EHR data. This model employs standardized vocabularies such as Systematized Nomenclature of Medicine-Clinical Terminology (SNOMED CT), Current Procedural Terminology (CPT), and the ICD (International Classification of Diseases) versions 9 and 10 for consistent data mapping. The deidentified version of the MU CDM, updated in October 2022 with altered dates and pseudo-identifiers, served as the foundation for our study. Database queries were executed using the Snowflake computing platform. The MU Institutional Review Board (IRB) approved this research.

#### Cohort identification

We utilized the framework from Furmanchuk (2021)^37^ to identify the diabetes mellitus (DM) cohort from our MU CDM based on the SUrveillance, PREvention, and ManagEment of Diabetes Mellitus (SUPREME-DM) algorithm. the SUPREME-DM DataLink is one example of a distributed registry developed for studying *Any*-DM (mixed Type 1 DM and Type 2 DM codes) using a standardized data extraction approach based on diagnosis, labs, and medications^38–40^. Although SUPREME-DM has not focused on distinguishing adults with Type I DM vs. Type II DM, this algorithm has been shown to have the potential of extracting the most representative EHR-based DM cohort^40^. We defined the MU study denominator as any patient between ages 18 to 89 years at the visit with at least two distinct encounter days. The encounter types included ambulatory visit (AV), emergency department (ED), emergency department admit to inpatient hospital stay (EI), inpatient hospital stays (IP), non-acute institutional stay (IS), and telehealth (TH) between 01/01/2010 and 01/31/2023.

We implemented the definition of SUPREME-DM on the MU denominator using the following steps (as detailed in Figure 2A): <u>Exclusion based on periods of pregnancy</u>. We first excluded pregnancy-related encounters using relevant ICD and CPT codes, then masked encounters within a year of each identified pregnancy. <u>Diagnosis codes</u>. We identified diabetic patients as those having two visits with diabetes-related ICD codes on separate days within two years, noting the date of the initial visit. <u>Lab codes</u>. Using LOINC IDs we filtered lab tests for HbA1c and glucose levels, identifying diabetic cases by two separate tests within two years, recording the date of the first. <u>Medications</u>. We identified diabetic patients through prescriptions for specific DM medications or non-specific medications when accompanied by a relevant diagnosis or lab test within two years. Finally, we combined data from diagnoses, labs, and medications to form the DM cohort, marking the earliest event date as an estimate for the DM onset. If a patient included a DM diagnosis in their first encounter, we marked that date as the DM onset estimate.

**Figure 2.**
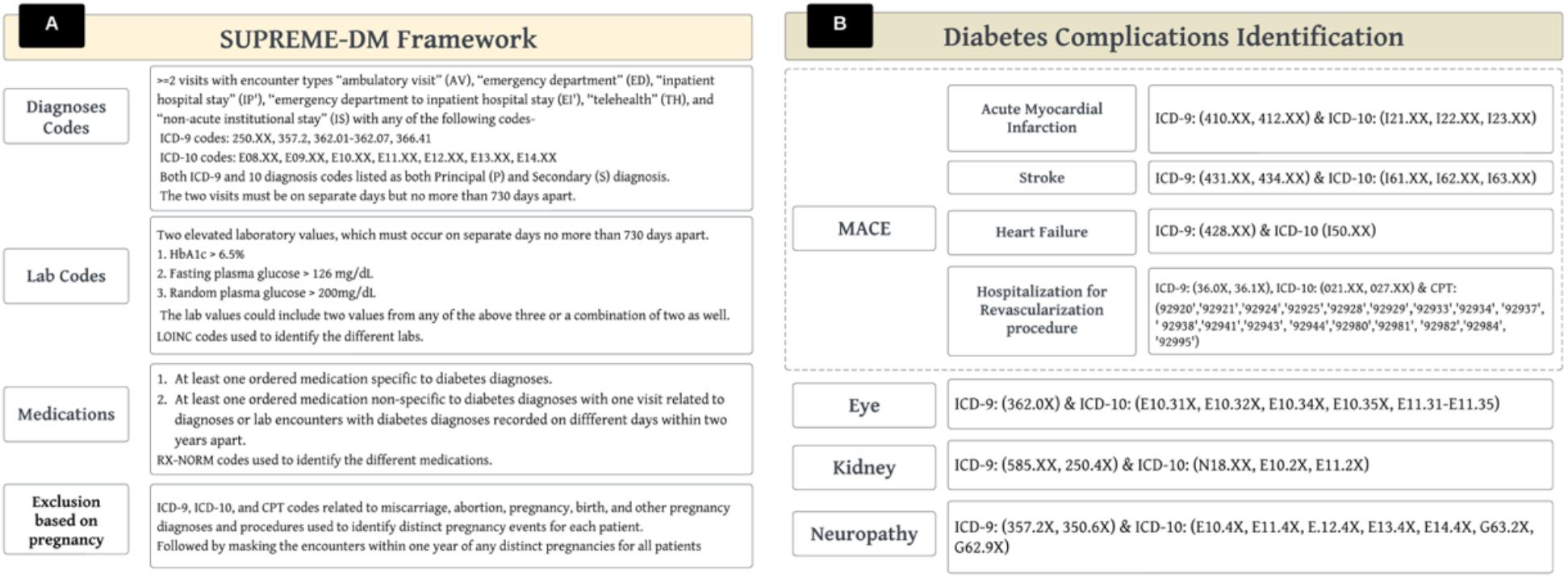
(A) shows the framework for constructing our diabetes cohort using SUPREME-DM from the MU EHR database. The process includes the exclusion of pregnancy-related encounters, identification of diabetes via ICD-coded visits and lab tests for HbA1c and glucose, and diabetic classification through specific medication prescriptions. The earliest diabetes indicator among diagnoses, lab results, or medications is marked as the estimated onset date for diabetes. Figure 2(B) illustrates the methodology for identifying major diabetic complications within the patient cohort. This includes the utilization of specific diagnosis and procedure codes to identify occurrences of retinopathy, chronic kidney disease, neuropathy, and MACE, with the latter encompassing myocardial infarction, stroke, heart failure, and revascularization hospitalizations. The initial encounter date for each complication is captured and recorded as the endpoint event date in the EHR data.

Figure 2B shows the diagnosis and procedure codes used to identify the four major complications among the diabetes patient cohort, including retinopathy, kidney disease (nephropathy), nerve damage (neuropathy), and major adverse cardiovascular events (MACE). The MACE events are acute myocardial infarction, stroke, heart failure, and hospitalization for revascularization procedures. For any of the four major complications identified, we recorded the date of the first recorded encounter for each complication in the EHR as the endpoint event date.

#### Outcomes, features, and study timeline

Figure 3-[1] shows how we structured a learning period for a hypothetical patient timeline in our EHR. This learning period determines the cutoff point for each patient in the identified DM cohort. The learning period for patients with any of the four diabetic complications spanned from their first EHR encounter up to the visit immediately preceding their first recorded complication event. For those without complications, the learning period extended to their last EHR encounter i.e. their learning period includes the entire EHR sequence. We also excluded patients whose diabetes diagnosis was recorded after their complication diagnosis to ensure diabetes was reported before the complication occurrences in the EHR timeline. The patient cohort derived from this process (cohort A) was used for pretraining. For finetuning and our Top-BERT experiments, we refined the cohort (cohort B) to only include patients with at least 5 but no more than 100 recorded encounters on different dates and with a minimum of five different diagnosis codes.

**Figure 3.**
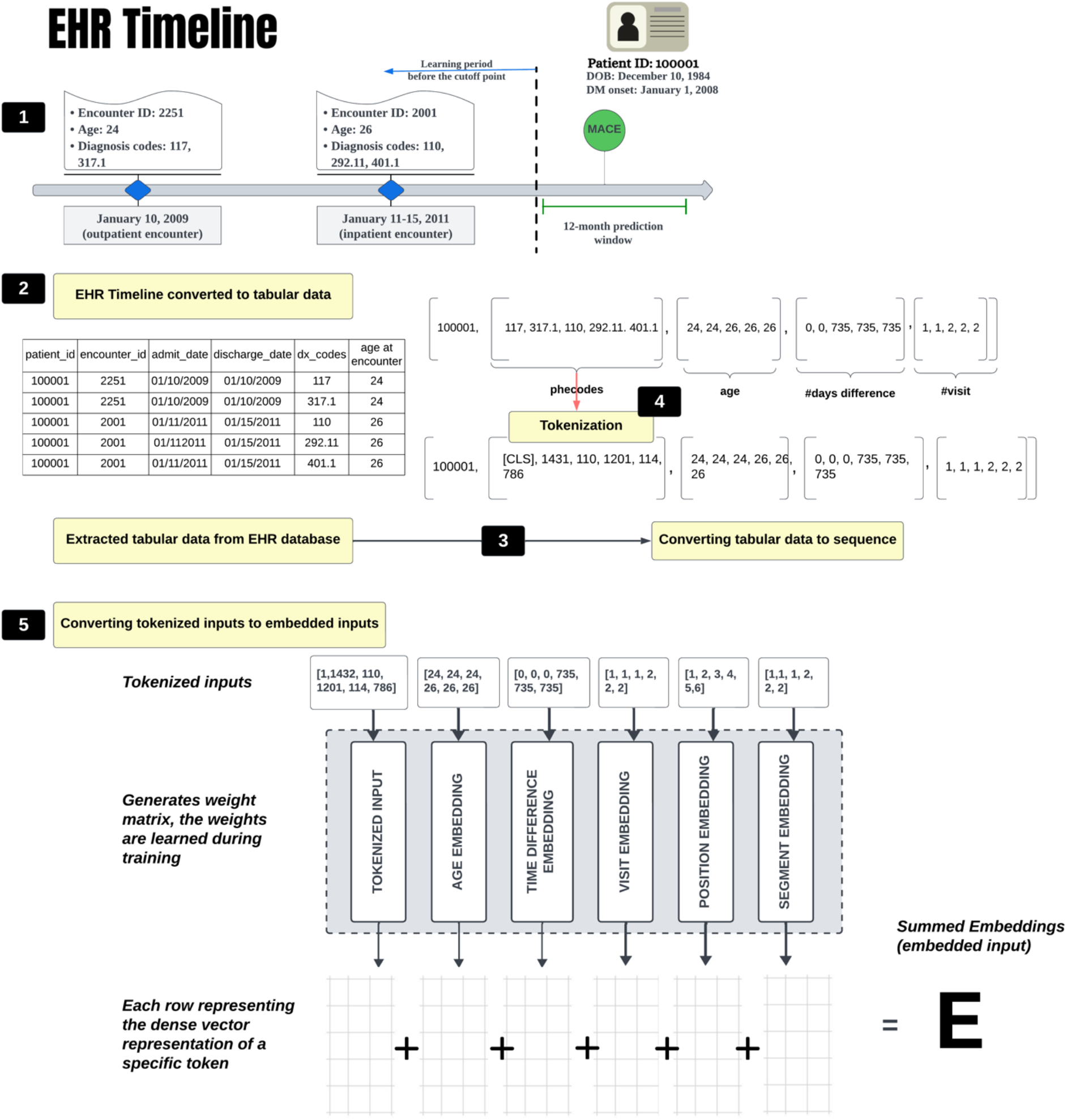
This figure illustrates [1] the methodology for structuring a learning period based on EHR data for patients with diabetes, defining the time frame leading up to either the earliest complication event or the last recorded encounter. The delineation of this period guided the selection of patient cohorts for pretraining and finetuning, with specific inclusion criteria based on encounter and diagnosis code counts to ensure relevance and accuracy in predictive modeling. The incidence of complications was identified in a designated prediction window following the learning period, contributing to the creation of outcome vectors for sequence classification tasks. This approach underpins the preprocessing of features for BERT, involving [2] the conversion of the timeline to structured tabular data, [3] the conversion of structured tabular data into sequential formats, and [4] the development of a tailored dictionary for EHR-specific tokenization,[5] culminating in the embedded input sequences that drive the training of the BERT-based model.

We defined the incidence for each complication (CKD, MACE, NEUR, and RET) within the first 12 months from the end of the learning period (prediction window). For instance, if a patient is diagnosed with MACE within the next 12 months, as shown in Figure 3-[1], after the end of its learning period, then the outcome is 1 else 0. Each patient can have multiple complications diagnosed in the prediction window. Thus, in the DM cohort, each unique patient ID is linked to four distinct labels, each binary and not mutually exclusive, to be used for our sequence classification task. For the binary classification task, we identified a common medical issue––prolonged hospital stays––assigning a value of 1 if the length of stay exceeded 7 days at any time during the EHR record of each patient. We extracted the labels of the four micro- and macro-vascular complications and prolonged hospital stay for all patients in the finetuning cohort B as our outcomes for the study. We curated separate datasets for pretraining and finetuning (cohorts A and B, respectively), extracting all encounters with diagnosis codes with admit-discharge dates and date of birth (DOB) within their specified learning period. The diagnosis ICD 9/10 codes are mapped to Phecodes^41^ to reduce sequence dimensionality. The DOBs were used to calculate the age at each encounter date. Finally, we constructed a tabular representation of the EHR timeline for each patient, as shown in Figure 3-[2].

#### Feature preprocessing for BERT

In this study, we used both diagnoses lists and temporal features––age at visit, number of visits, and inter-visit time difference in days––as integral components of the input to the embedding layer within our BERT-based model. To facilitate the training of BERT with EHR data, we initially converted the structured tabular data into a sequential format, as depicted in Figure 3-[3]. This entailed creating the temporal attributes as sequences, aligning each diagnosis code (in order of occurrence) to corresponding temporal attributes, and standardizing the sequence length for each patient’s record. Acknowledging the distinct nature of EHR diagnosis codes as opposed to traditional text, we constructed a unique dictionary. This lexicon assigns a discrete numerical identifier to each diagnosis code and each special token, such as [CLS], [PAD], and [MASK], thus creating a bespoke mapping system tailored for EHR data. Subsequently, we employed this dictionary for tokenization, transforming the list of Phecodes into a sequence of tokens with a [CLS] token inserted at the beginning, as illustrated in Figure 3-[4]. These tokenized sequences are what we leveraged as the input features for the embedding layer. Figure 3-[5] shows the procedure for formulating the embedded input sequences essential for training our BERT-based model. Each tokenized input sequence with its corresponding temporal attributes is used as input for the embedding layer. This layer is responsible for transforming each token into a dense vector representation, generating weight matrices for each layer of the model. The dimensions of the matrices are determined by the sequence’s length and hidden size parameter of the model. Finally, the summed embeddings are fed into the model’s subsequent layer to complete the training process.

### Experimental Design

#### Model training details and experiments

Our experimental design involved two distinct cohorts: an unlabeled cohort for pretraining (cohort A) and a labeled cohort (cohort B) for finetuning, as well as training our Top-BERT model. We randomly allocated cohort B into training, validation, and test sets in a 7:1:2 ratio for use in both finetuning and Top-BERT training. We conducted experiments to train, test, and evaluate the Top-BERT architecture on predicting four major micro and macro-vascular complications–– CKD, MACE, NEUR, and RET–– and prolonged hospital stays. Furthermore, we developed a pretrained model using cohort A and finetuned the model using cohort B to predict these micro and macro-vascular complications. We implemented a PyTorch^42^ workflow to run our BERT-based experiments. Additionally, we benchmarked our model against the popular machine learning model XGBoost^43^.

##### Top-BERT training and evaluation

We trained our Top-BERT model using three tasks simultaneously, namely, mask language model (MLM), binary classification for prolonged hospital stay, and a sequence classification for the 12-month prediction of the four major diabetic complications. For the MLM task, we masked 15% of the tokens in each input sequence, following a strategy where 80% of the masked tokens were replaced with a special [MASK] token (denoted as -1), 10% were replaced with a random token from the vocabulary, and the remaining 10% were left unchanged. The masked tokens were augmented with the processed input features to train the model. This masking strategy introduces noise and variability into the input data, encouraging the model to learn robust and context-dependent representations of the EHR data. The primary objective of the MLM task is to accurately predict the original tokens at the masked positions. During training, a cross-entropy loss function is utilized, which ignores the masked tokens to concentrate on the contextual learning of unmasked tokens. For evaluation, the model’s performance is exclusively assessed on its capacity to predict the masked positions correctly.

Simultaneously, the binary task tokens were generated, indicating prolonged hospital stays (greater than 7 days) during the input feature-augmentation process for model training. For the prediction process, the logits from the model’s output —derived from the pooled output and, subsequently, a dense layer— are processed through a softmax layer to calculate predictive probabilities. Binary cross-entropy loss function was used to minimize the prediction error. The third task, focused on sequence classification, predicts four types of micro and macro-vascular complications (CKD, MACE, NEUR, and RET). In this component, the model channels the pooled output through a dense layer to generate logits. These logits are then transformed into probabilities using the sigmoid activation function. The model is fine-tuned with a weighted binary cross-entropy loss function to optimize the predictive accuracy. The combined loss from all three tasks is backpropagated through the layers to learn the weights. We used the AUROC metric to evaluate model performance as the primary evaluation metric like previously established frameworks^27,28^. We employed the best-performing trained model for evaluation against our test data and report all results on it.

##### Pretraining-finetuning

During the pretraining phase, we employed conventional MLM and binary tasks (prolonged hospital stay) utilizing cohort A. Post-training, the model demonstrating optimal performance was further finetuned on cohort B. Loading the pretrained weights, two finetuning variations were employed, each utilizing different encoder output strategies. Finetuning-A employed conventional [CLS] token embeddings. Finetuning-B innovated with a custom prediction head utilizing the last encoder layer’s embeddings refined through three dense layers and ReLU activations, with the final layer producing logits. We implemented a weighted random sampler during batch training to address class imbalance in the four label predictions.

##### Comparison with an ML model

XGBoost was employed as a time and context unaware benchmark for comparison with our time and context aware BERT-based models. We transformed the diagnosis codes into one-hot encoded features across our training, validation, and test datasets. Each Phecode was denoted by a binary flag, and we also included the age at the last visit as an additional feature. We utilized a multioutput strategy with XGBoost to predict the four complications simultaneously.

##### Metrics for model comparison

To assess the performances of the four classification tasks (CKD, MACE, NEUR, and RET) in BERT-based models and XGBoost, we computed micro-averaged AUROC (mAUROC). The mAUROC pools the individual true positives, false positives, false negatives, and true negatives across all classes and then computes the AUROC from these combined totals, which indicates the overall performances of the models in distinguishing between both majority and minority classes across various thresholds^44^. Additionally, to gain insights on the differences and similarities of Top-BERT and XGBoost in handling class imbalance, we compared the model performance for each classification task using precision, recall, F1-score, Mathew’s correlation coefficient (MCC) and confusion matrix computed at varying threshold values (10%, 30%, 50%, and 80%).

#### Temporal representation comparison using embeddings

In our investigation, we explored the impact of temporal factors—patient age, visit order, and inter-visit time differences—when integrated into the embedding process of predictive models for diabetic complications. We conducted ablation studies to comprehensively evaluate the utility of various temporal embeddings.

In our Top-BERT experiments, we compared 15 distinct models, each featuring a distinct embedding layer architecture that integrated various temporal components. The distinct models we compared also include the three pioneering frameworks of BERT in EHR data– BEHRT^21^ and Med-BERT^27^. The mAUROC metric served as our primary performance indicator, reflecting each model’s discriminative power in predicting diabetic complications. Concurrently, we utilized Shannon’s entropy^32^—a measure of the unpredictability or complexity of information content—to gauge the informativeness of the embeddings generated by each model. We obtained the embeddings from the final encoder layer for each model, transformed them into probabilities via softmax, and then calculated their total entropy using the formula *H*(*X*) = − ∑_*x* ∈X_ *p*(*x*) log *p*(*x*). A higher Shannon’s entropy value indicates a richer, more complex embedding representation, suggesting that the model captures a greater amount of information from the input data. Information gain was calculated by comparing the entropy values of each model to that of a comprehensive model inclusive of all temporal factors–– incorporating input, visit number, inter-visit time difference, age, and positional and segmental embeddings––providing a metric for the relative improvement in predictive power.

Furthermore, we performed ablation studies across different embedding layer architectures within our pretraining and finetuning workflow, creating 15 unique pretrained models. Subsequently, we executed 15 finetuning experiments each for scenarios A and B. By juxtaposing mAUROC scores with information gain values; we aimed not only to identify the model with the highest predictive accuracy but also to discern which embedding design best encapsulates the complexity of patient history in our single-center dataset. This dual assessment allowed us to balance the trade-off between model simplicity and the depth of temporal understanding necessary to accurately depict the progression towards diabetic complications.

#### Model Explanations

We have tailored the Integrated Gradients^45^ methodology for application on our Top-BERT model to discern feature-level attributions that influence our predictive models on both an individual patient and a global level. Integrated Gradients is a feature attribution method that assigns an importance score to each input feature of a neural network by integrating the gradients of the model’s output with respect to the input features, tracing a path from a given baseline to the actual input. This method is designed to satisfy two fundamental axioms: sensitivity and implementation invariance, ensuring reliable and consistent attributions.

Formally, for a given input *x* and a baseline *x*′, the integrated gradient along the *i*-th dimension is defined as:

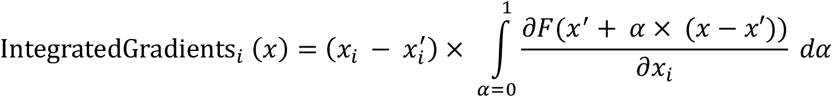

Here, *F*: ℝ^*n*^ → [0,1] is the model, *x* is the input, *x*′ is the baseline, *x* is the *i*-th feature of *x*, and 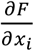 is the gradient of *F* with respect to *x*_*i*_. The integral accumulates the gradient for feature *i* at all points interpolated between the baseline and the input. A practical baseline for language models is the all-zero input embedding vector. We approximate the integral via a summation called the Reimann approximation. To apply Integrated Gradients to Top-BERT, we consider the model function *F*(*x*) as the output logit resulting from the input sequence *x* after transformation through the model’s embedding and subsequent BERT layers: *F*(*x*) → *Head* (*BertLayers*1*EmbeddingLayer*(*x*))), which is ***h***_*seq*_ for Top-BERT. We divide the linear path between the baseline and the input into α increments to calculate the gradients at each step, which are then aggregated and normalized to determine the contribution of each feature to the prediction for each outcome. Additionally, to identify the global-level feature attributions, we averaged attribution scores for each feature in each patient and then calculated a sum over these feature-level attributions across all patients for each outcome. Furthermore, we computed mean attribution scores within distinct age groups to identify age-specific patterns of feature importance for each outcome, thereby enhancing the explainability and transparency of our model in the clinical domain.

##### Algorithm 1. Adaptation of Integral Gradients Algorithm for Sequence Classification using Top-BERT Input

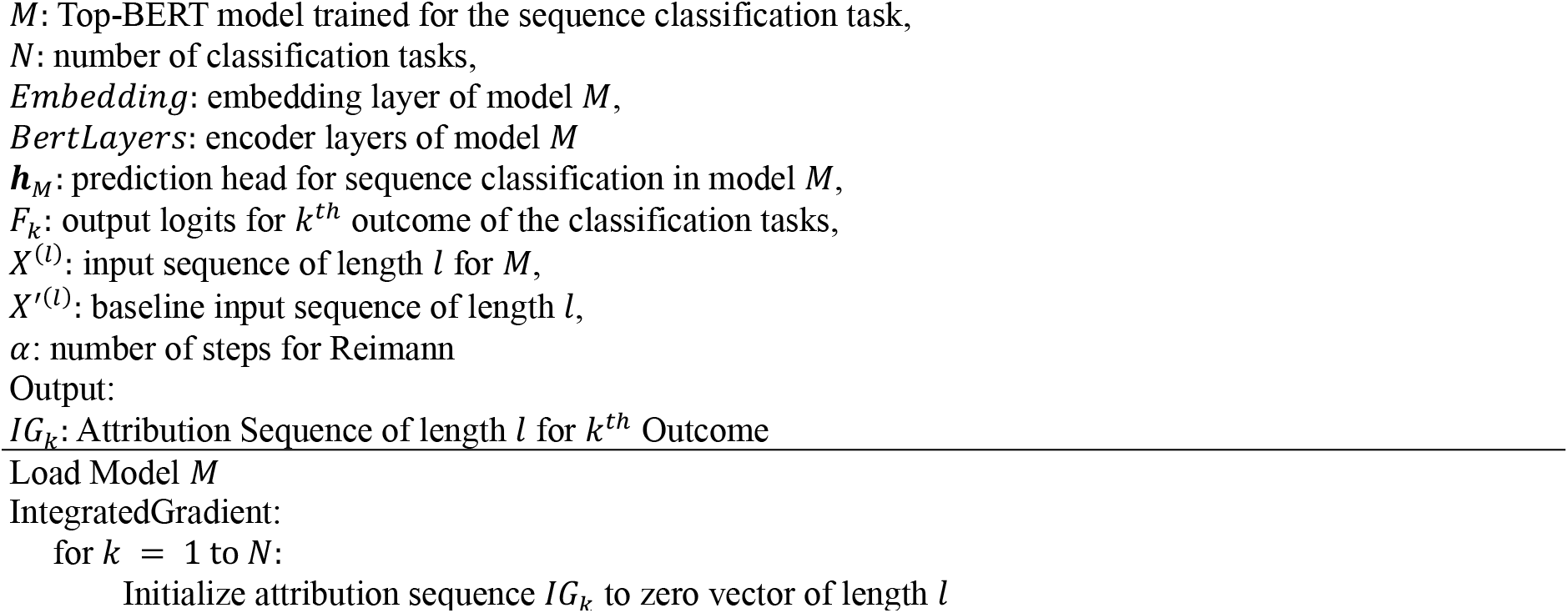
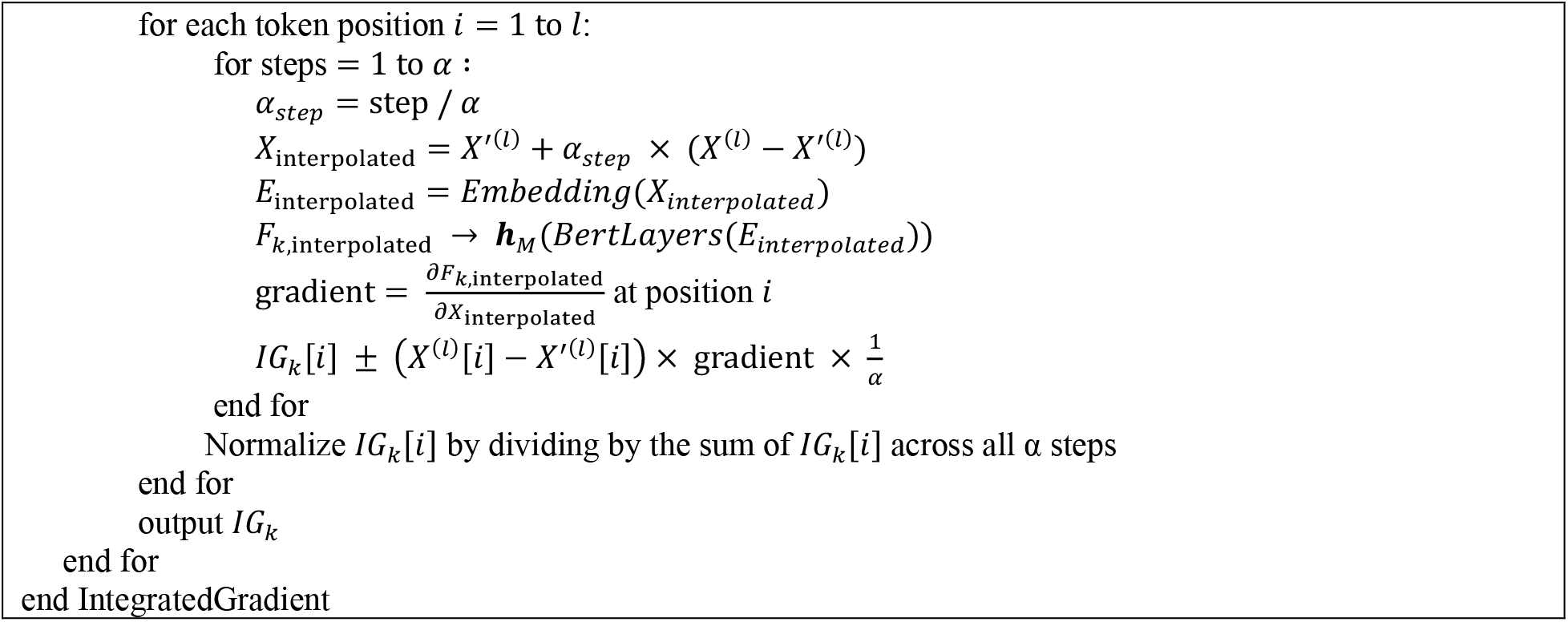

## Results

### Data summary

Figure 4 provides a detailed demographic and clinical profile of the diabetes patient cohort used in this study. Table 4A contrasts two subsets: the initial pretraining group (Cohort A), comprising 50,993 patients, and the subsequent finetuning/Top-BERT group (Cohort B), which includes 36,539 patients. The average number of visits per patient for pretraining cohort A (∼31 visits) is higher than cohort B (∼27 visits). Both subsets exhibit similar averages in number of diagnoses per patient (∼44 diagnoses per patient). The average age for both cohorts is about 60 years and primarily white (∼89% white), the cohorts maintain a consistent gender ratio. Prolonged length of stay was approximately 19.4% for both cohorts. Figures 4B through 4E show further insights into Cohort B, shedding light on healthcare interaction patterns and the prevalence of specific health complications within this group. Specifically, Table 4D indicates high occurrences of hypertension (69%), hyperlipidemia (51.7%), obesity (34.1%), and tobacco use disorder (33.53%) within the cohort, which are predominantly categorized under endocrine/metabolic and circulatory system disorders. Furthermore, approximately 10.7% of patients had neuropathy, 10.9% had MACE, 7.4% had CKD, and 2.7% had retinopathy.

**Figure 4:**
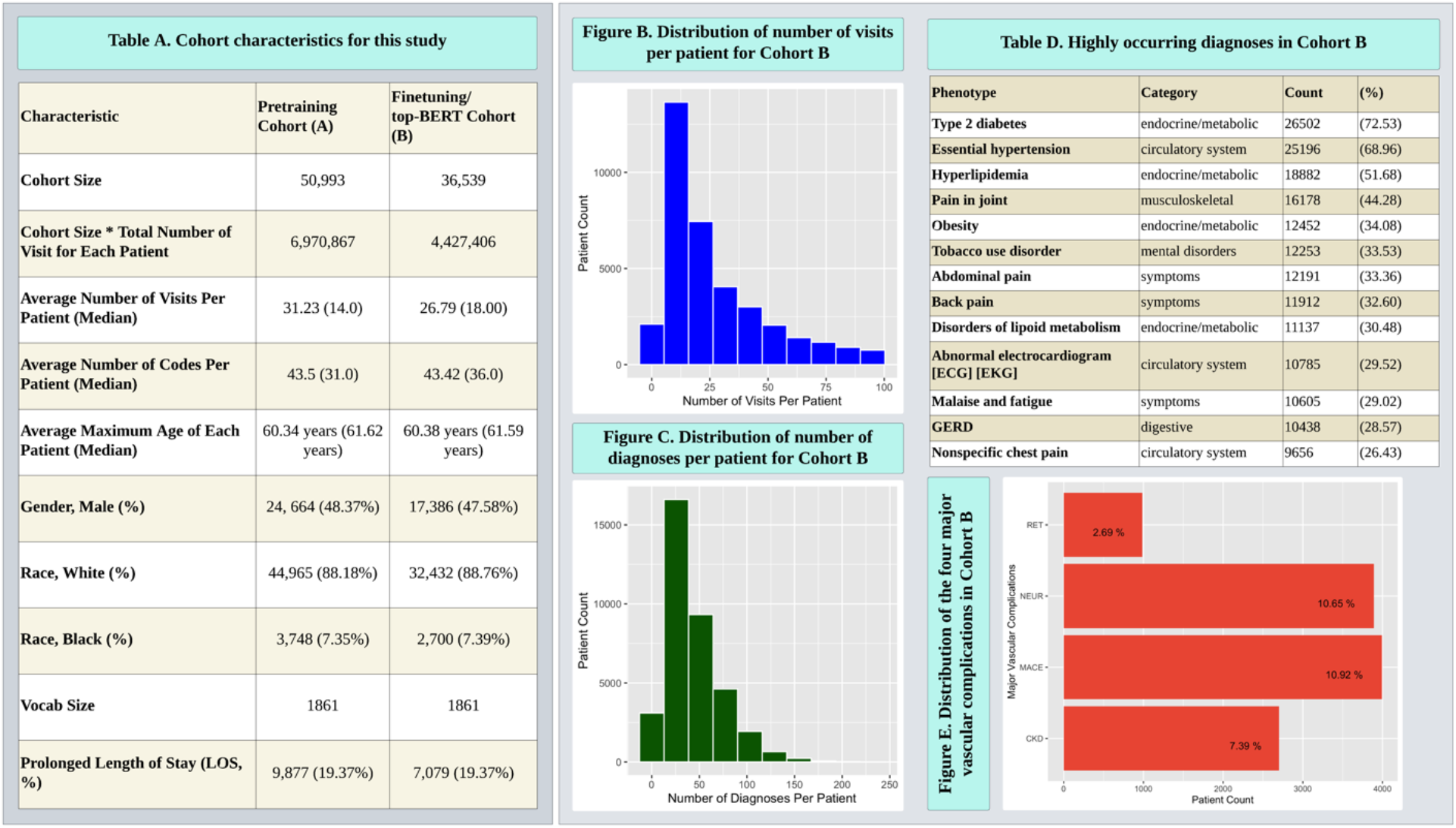
This figure provides a comprehensive summary of the cohort’s demographic and health status, the distribution of healthcare interactions, and the prevalence of significant health conditions, which are critical for the subsequent analysis of the study. Table 4A outlines the characteristics of the study cohorts, revealing a pretraining cohort (A) size of 50,993 and a finetuning/Top-BERT cohort (B) size of 36,539. Figure B illustrates the distribution of the number of visits per patient for cohort 4B, which shows a right-skewed distribution, indicating that most patients have fewer visits, with the number tapering off as the visit number increases. Figure C depicts the distribution of the number of diagnoses per patient for cohort B, which is also right-skewed, with more patients having diagnoses number in the mid-range of 100. Table 4D lists the most frequently occurring diagnoses in cohort B, with type 2 diabetes, essential hypertension, and hyperlipidemia being the most common. These conditions are predominantly categorized under endocrine/metabolic and circulatory system disorders, reflecting the health concerns prevalent in the cohort. Figure E depicts the relative distribution of the four primary study outcomes, indicating a higher frequency of neuropathy and major adverse cardiovascular events (MACE) within the patient cohort.

### Model Performance

In evaluating the performances of the three distinct architectures for predicting four major complications within Cohort B, our Top-BERT model outperformed the finetuning adaptations of pretrained models and the time and context unaware approach using XGBoost. Our test data consisted of 7.25% of CKD labels, 11.6% of MACE, 10.9% of NEUR, and 2.7% of RET. As shown in Figure 5A, the highest mAUC was achieved by the Top-BERT design input+visit+time_diff (mAUC of 0.7125), while the same embedding configuration in the finetuning experiments had lower mAUCs (0.6786 and 0.689, respectively). In contrast, XGBoost yielded an mAUC of 0.5208. Moreover, Figure 5C highlights the ROC of the best-performing model configuration (input+visit+time_diff) with the finetuning frameworks at various thresholds.

**Figure 5.**
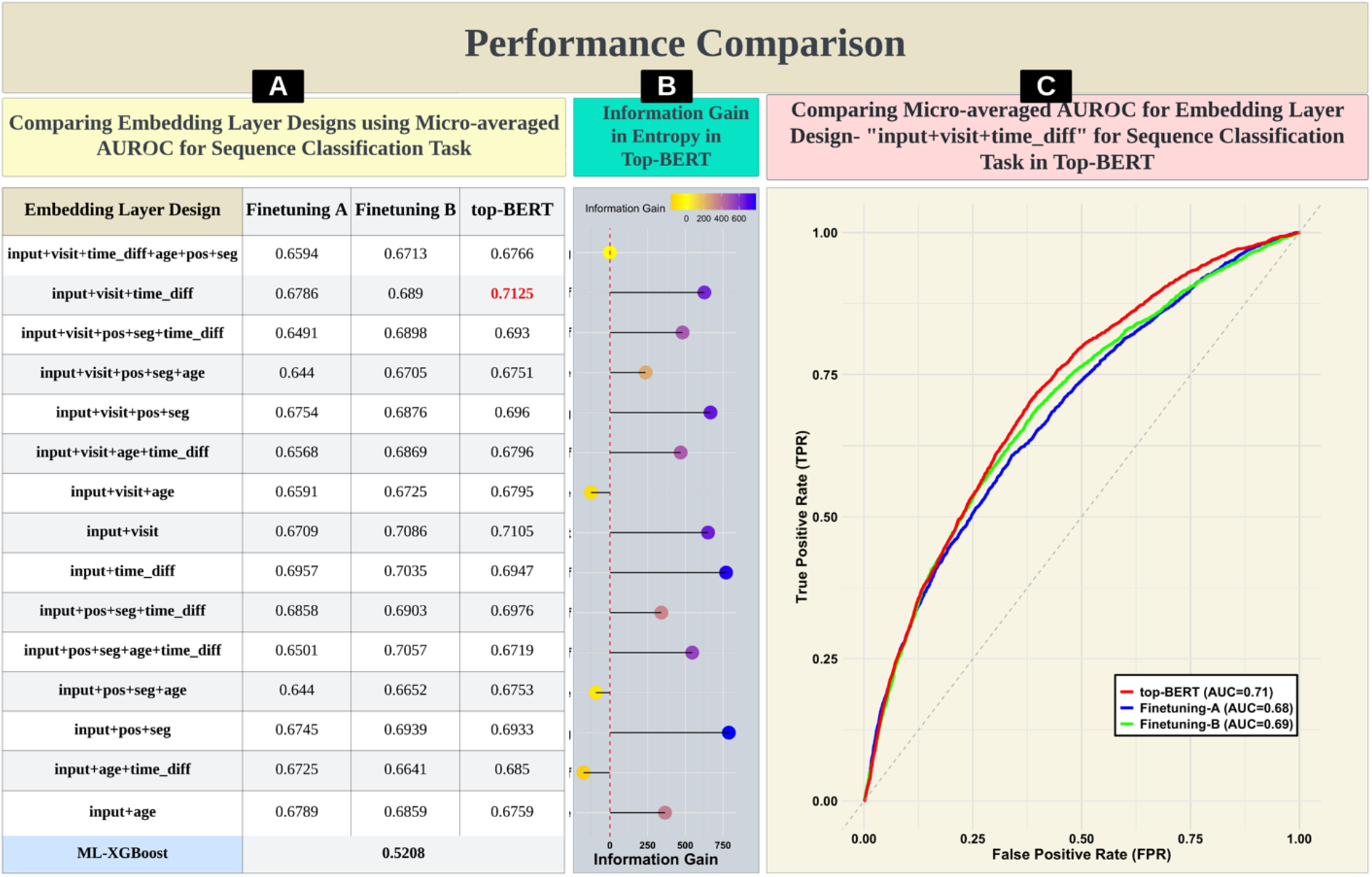
This figure shows a comprehensive depiction of the performance comparison of our proposed Top-BERT framework with the two finetuning frameworks adapted for this study for predicting the four major complications within Cohort B. Figure 5A displays a table comparing various embedding layer designs based on micro-averaged AUROC scores for three different model training scenarios: Finetuning A, Finetuning B, and the final Top-BERT model. The highest-performing design in the Top-BERT model is highlighted, suggesting it has the best trade-off between complexity and performance. Figure 5B visualizes the information gain in entropy for the Top-BERT model across different embedding designs. Each point represents a different design, with its position on the x-axis indicating the amount of information gain using the most complex model (input+visit+time_diff+age+pos+seg) as a baseline for comparing the gain. This graph helps assess which design captures the most relevant information from the data. Figure 5C compares the micro-averaged AUROC curves of the Top-BERT model and the two finetuning phases, illustrating the true positive rate (TPR) against the false positive rate (FPR) for highest-performing design. The area under the curve (AUC) for each model is annotated, allowing for a direct comparison of their predictive performance.

We observed (Figure 5B) that an embedding incorporating input features with temporal elements, such as visit number and inter-visit time difference, alongside positional and segmental embeddings, yielded a higher information gain than the more complex, feature-rich embedding design (input+visit+time_diff+age+pos+seg). Within Top-BERT, the embedding approach utilized in BEHRT (input+age+pos+seg)^21^ demonstrated a lower information gain, whereas the Med-BERT (input+visit)^27^ embedding configuration showed higher information gain, while with AUCs of 0.6753 and 0.7105, respectively.

Table S2 in the supplementary document compares the five Top-BERT embedding configurations (having comparable mAUROCs) and XGBoost using various metrics at varying thresholds to gain insights on their differences and similarities in handling class imbalance. We observed that higher precision values and lower recall values obtained by XGBoost were consequences of conservatively predicting fewer number of positive cases (both true positives and false positives) in all classification tasks. On the other hand, Top-BERT showed better measure of separability, as indicated by higher mAUROC values, and higher efficiency in accurately predicting the positive classes compared to XGBoost (details in supplementary document).

### Model Explanations

Figure 6 highlights the aggregated global-level feature importance for the four outcomes of this study: CKD, MACE, NEUR, and RET. In Figure 6, the left panel shows the common diagnoses predictive of all four outcomes, while the right panel shows the significant features contributing (positively) to each outcome separately. In the figure, the color intensity representing the attribution score for each diagnosis reflects the combined influence of the average attribution score per patient and the prevalence of that diagnosis within each respective outcome cohort.

**Figure 6.**
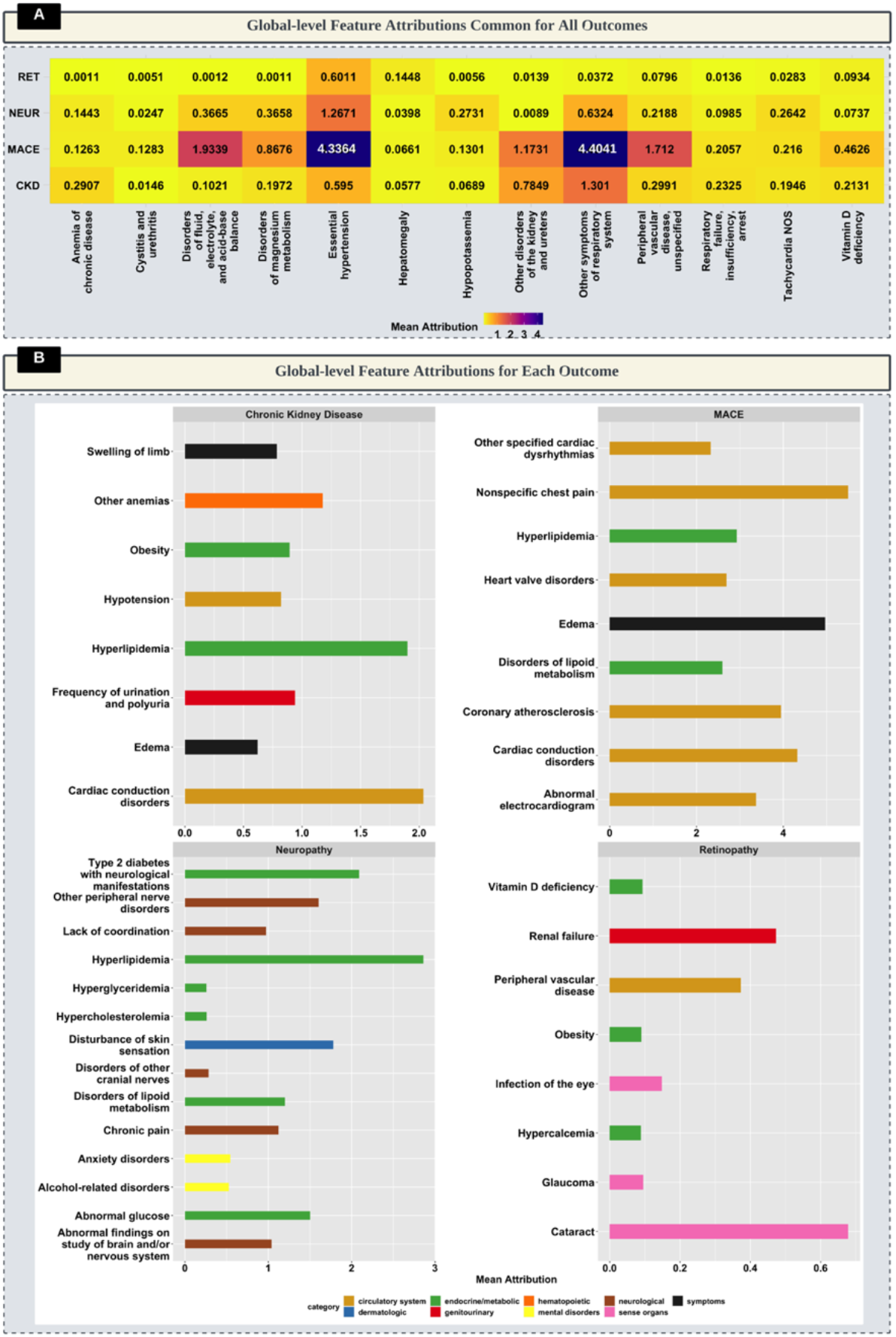
Global-level feature attributions for four major diabetes-related complications. The top panel (A) shows the common feature attributions across all outcomes, while the bottom panel (B) details the top contributing features for each specific complication: chronic kidney disease (CKD), Major Adverse Cardiac Events (MACE), Neuropathy (NEUR), and Retinopathy (RET). In Figure 6A, each column represents a diagnosis with its corresponding aggregated mean attribution score, represented in color-coded heatmaps, indicating the strength of prediction with the complication outcomes. In Figure 6B, each row is the aggregated mean scores for the diagnosis and categories of diagnoses are color-coded, as indicated in the legend, facilitating a comparative visualization of feature importance across the different diagnostic classifications. Note that the mean attribution scales vary for each complication to optimize the visual representation of the data, facilitating the comparison of influential predictive diagnoses within each respective outcome.

### Common Predictive Features

Metabolic abnormalities such as vitamin-D deficiency, hypopotassemia, magnesium metabolism disorders, and disorders of fluid, electrolyte, and acid-base balance emerged as common important predictors for all studied outcomes. Additionally, circulatory system conditions, including hypertension, tachycardia, and peripheral vascular disease, alongside respiratory system disorders and hematopoietic conditions, such as chronic anemia, were found as shared predictive features for increased risk of all four outcomes (Figure 6, left panel).

### Outcome-Specific Feature Importance

Edema and cardiac conduction disorders highly indicated increased risk for CKD and MACE. Similarly, disorders of lipoid metabolism had high attribution for predicting both MACE and NEUR. Hyperlipidemia had high attribution scores for CKD, MACE, and NEUR. Additionally, obesity was highly predictive for both CKD and RET.

Symptoms such as swelling of limbs, low blood pressure (hypotension), and urinary system conditions such as disorders of kidney/ureter and frequency of urination were found to be notable for their contribution to CKD risk. For MACE, various circulatory system disorders such as abnormal electrocardiogram, coronary atherosclerosis, chest pain, cardiac dysrhythmias, and heart valve disorders– were identified as critical indicators. NEUR risk was closely linked with early neurological manifestations from Type 2 diabetes, peripheral nerve disorders, coordination issues, and chronic pain accompanied by abnormal findings related to the brain or nervous system. Interestingly, mental health disorders, including anxiety and alcohol-related disorders, also had high attribution for predicting NEUR. Furthermore, metabolic conditions such as hypercalcemia with early onset of eye-related symptoms such as infection, glaucoma, cataracts attributed significantly to increased risk of RET.

### Age-specific Feature Importance

Figure 7 shows the age-specific predictive features common across all outcomes (each diagnosis was present in at least one age group) of this study. Hypertension was shown to be a predictive factor for all outcomes for all age groups. In the 18-35 age group, the presence of hypertension, hyperlipidemia, and obesity attributed to an increased risk for all four outcomes. For individuals aged 35-55, abnormal glucose levels, alcoholism, hypoglycemia, and respiratory disorders were highlighted as significant predictive indicators. Among those in the 55-75 age range, obesity, disorders of lipoid metabolism, and diabetes diagnoses stood out as key common predictors of high risk for all outcomes. In the population over 75, hyperlipidemia and the prolonged use of anticoagulants were identified as common indicators across the studied outcomes.

**Figure 7.**
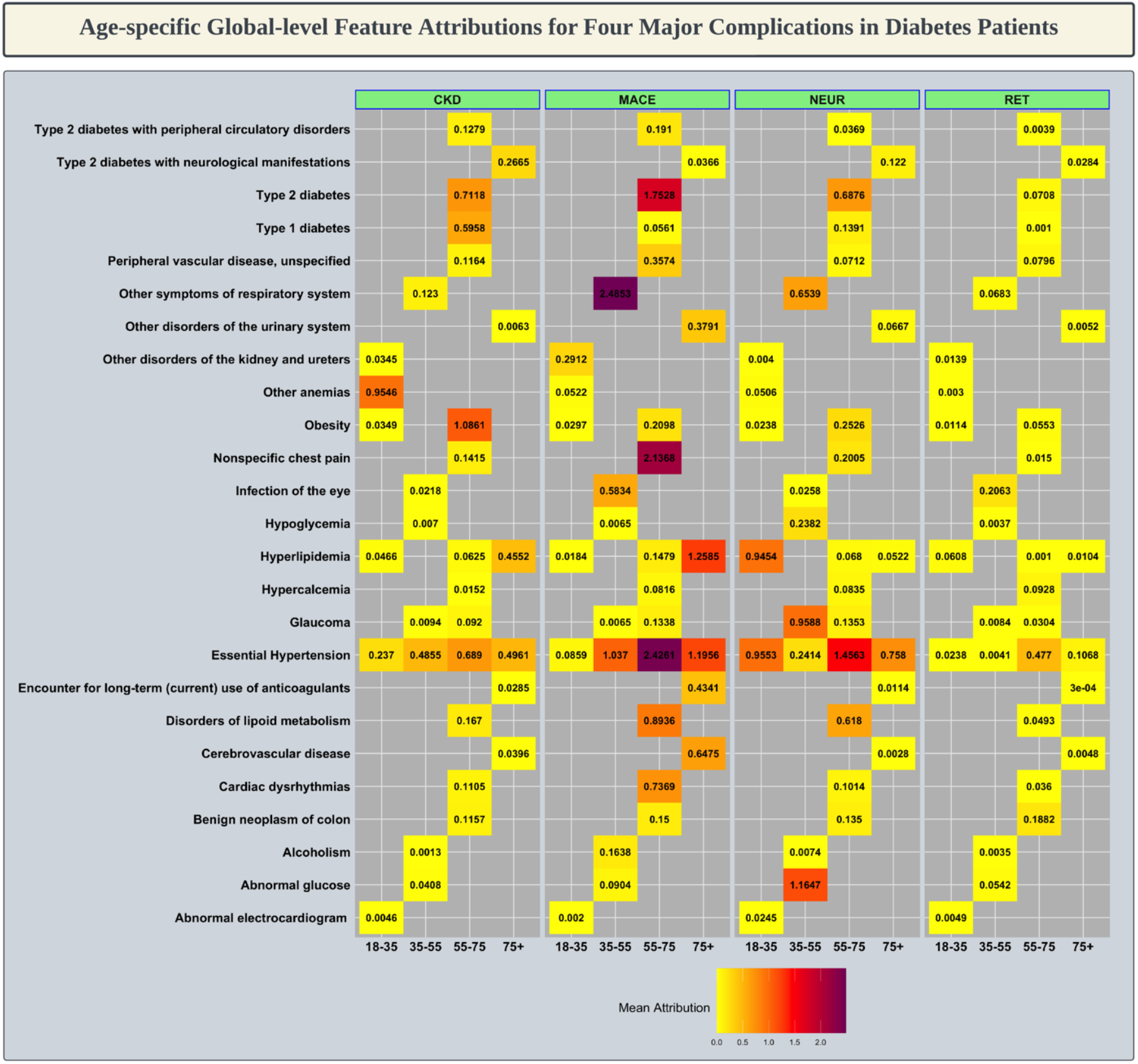
Heatmap illustrating age-specific global-level attributions of the common predictive features for four major complications in diabetes patients (each diagnosis is present in at least one age group). Each row represents a distinct diagnosis, while columns denote the patient age groups and the complications—chronic kidney disease (CKD), major adverse cardiovascular events (MACE), neuropathy (NEUR), and retinopathy (RET). Color intensity reflects the mean attribution value for each feature, with warmer colors indicating higher attribution (with higher incidence) and cooler colors indicating lower attribution, signifying the relative importance of each feature in the model’s predictions across different age brackets.

### Patient-specific Feature Importance

Figure 8 illustrates the attribution scores of various conditions diagnosed, computed using IG, for each visit recorded in the EHR. The figure provides a visual narrative of patient-specific features contributing to increased risk for the four outcomes examined, as determined for four randomly selected patients. For example, the timeline for a patient at elevated risk for Major Adverse Cardiac Events (MACE) (top-right quadrant) reveals atrial fibrillation and cardiac dysrhythmia as consistent predictive features across visits from November 2016 to January 2018, preceding a MACE diagnosis within the subsequent year. Notably, hyperlipidemia and hypertension emerged as indicative features of MACE risk in later visits. In another case, a patient at high risk for chronic kidney disease (CKD) (top-left quadrant) showed predictive features such as kidney or bladder cancer, along with hypertension, hyperlipidemia, and disorders of lipoid metabolism in visits leading up to the CKD diagnosis.

**Figure 8.**
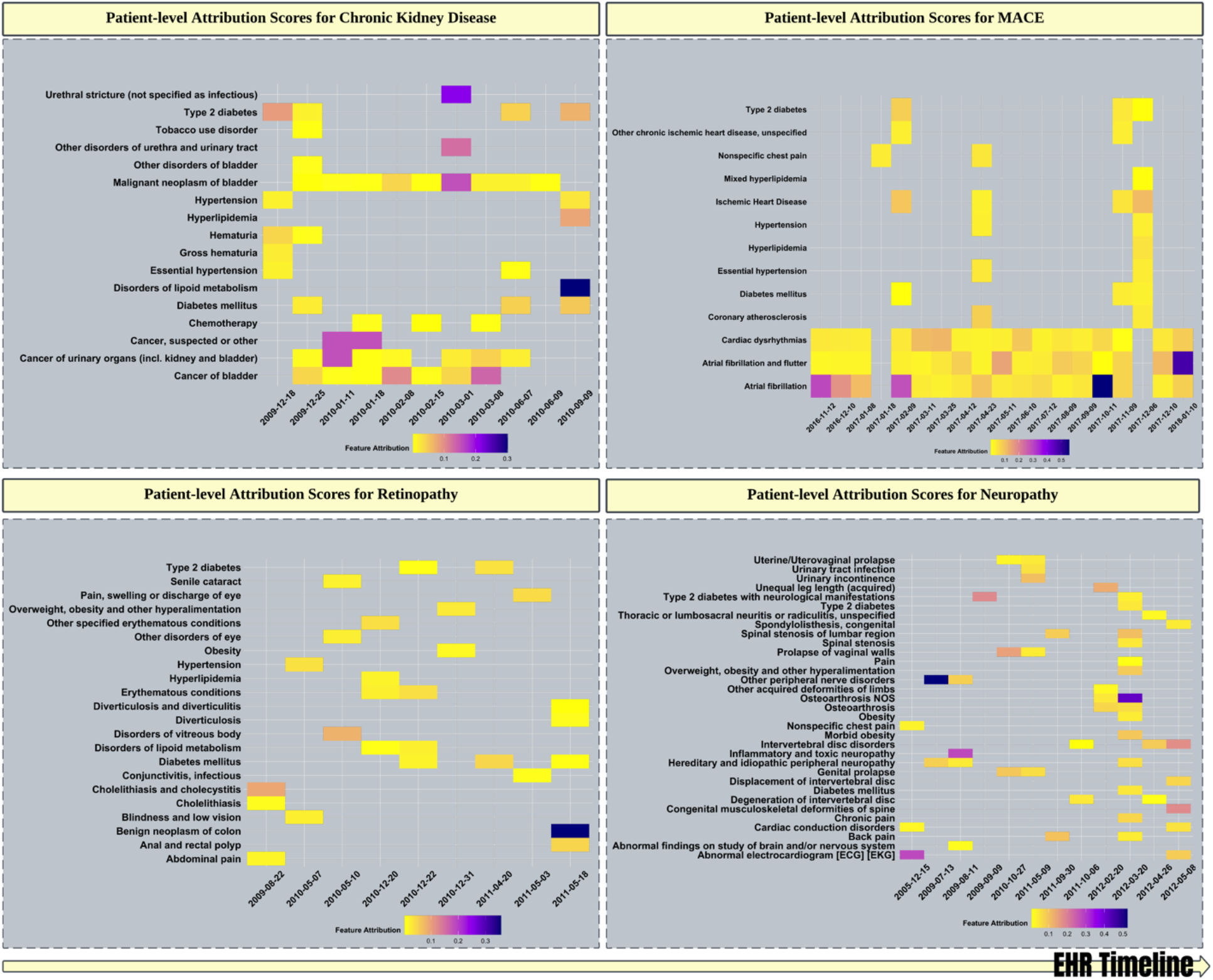
Heatmaps of patient-level attribution scores for four randomly chosen patients within our study cohort who developed major diabetic complications within a year following their last recorded visit. The visualizations present individualized feature attributions within each patient’s EHR timeline for chronic kidney disease (CKD), major adverse cardiovascular events (MACE), retinopathy, and neuropathy. Each row represents a specific diagnosis, while columns represent the encounter dates, showcasing how attribution scores vary throughout the patient’s medical history. Color gradients indicate the magnitude of the feature’s attribution to the respective complication, with warmer colors denoting higher attribution scores. This patient-centric analysis highlights the differential and time-related impact of various medical conditions on the risk of each complication.

### Implementation Details

Figure 9 outlines the computational framework employed in our study, utilizing Snowflake, AWS Workbench, the Nautilus HyperCluster, and PyTorch(2.0). Our process began with executing SQL queries within Snowflake for cohort identification from our EHR database. Subsequent data preprocessing and feature generation for model training were conducted within AWS’s secure environment.

**Figure 9.**
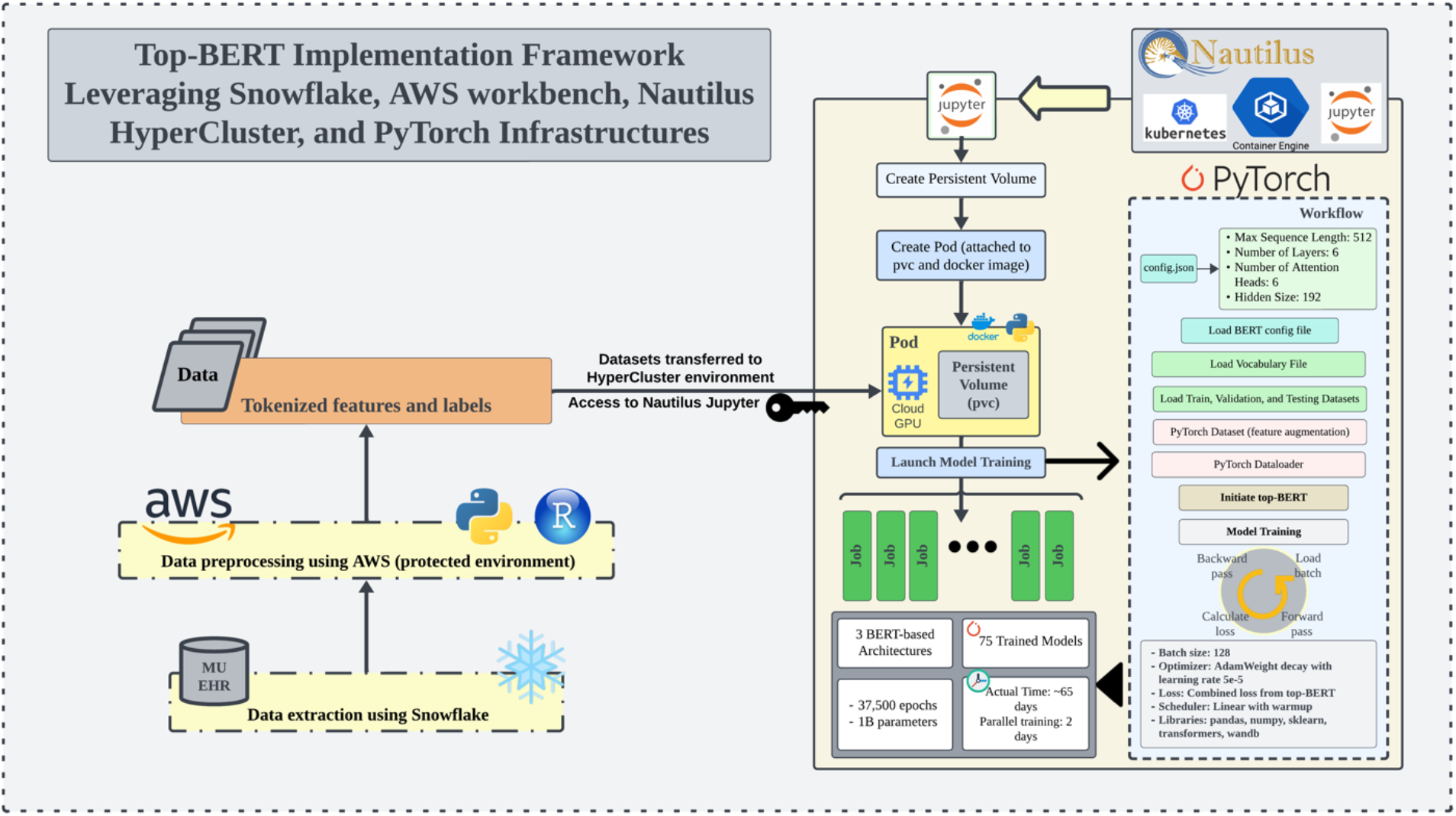
The diagram outlines the Top-BERT Implementation Framework, integrating various technological platforms for model development and training. The process begins with data extraction from MU EHR using Snowflake, then preprocessing in AWS’s secure environment. Tokenized features and labels are then transferred to the HyperCluster environment facilitated by Nautilus Jupyter to access GPU resources. Model training is initiated in this environment via persistent volumes and pods, leveraging Kubernetes orchestration. The PyTorch workflow incorporates configuration of BERT’s parameters, vocabulary loading, and dataset preparation for training 75 models across 3 BERT-based architectures. Training efficiency is exemplified by the reduction of actual training time to approximately 65 days, achieved in just 2 days due to parallel computing. This streamlined process underpins our study’s robust analytical capability.

The tokenized features and labels were transferred to the HyperCluster environment called NRP (National Research Platform) Nautilus, a nationwide cyberinfrastructure led by the Greater Plains Network. Figure 9 further illustrates the use of Jupyter IDE to implement model training across multiple experiments. We allocated persistent volumes to store datasets and orchestrated containerized jobs with the necessary docker image and GPU support. Specifically, NVIDIA A10 GPUs with 25 GB each were employed to facilitate parallel execution of tasks. Figure 9 also presents the workflow of our PyTorch-based implementation. It details the hyperparameters selected for our experiments and lays out the sequential steps in training the models. This systematic approach underpins the rigorous development and validation of our predictive models. We customized the Transformers library’s BERT model for model training to construct our unique Top-BERT architecture. Monitoring and managing training progress were achieved through Weights and Biases (wandb.ai). Using Nautilus’ parallel computing capabilities drastically reduced our total training duration for all experiments from an estimated 65 days to just 2 days, leading to significant efficiency gains.

Additionally, we implemented XGBoost to predict four micro and macro-vascular complications, utilizing the xgboost library, scikit learn’s MultiOutputClassifier, and the Optuna library for hyperparameter optimization. Model evaluation metrics were computed using scikit learn.

## Discussion

### Methodological and Technical Advancement

Our study demonstrates a notable advancement in the application of BERT’s architecture for clinical predictions from EHR data, particularly when constrained by the sample size typical of single-center datasets. Through the innovative implementation of task-oriented predictive (top)-BERT, we have demonstrated the adaptability and strength of the BERT architecture in facilitating an end-to-end training and evaluation approach. Top-BERT, utilizing the sequential input structure, embedding layer, and encoder stacks inherent to BERT adopts a multitask approach that integrates the conventional Masked Language Model (MLM), a binary classification for prolonged hospital stays, and multilabel sequence classification for micro and macro-vascular complications in diabetic patients. Our findings showed that Top-BERT can outperform both standard pretraining-finetuning BERT applications and traditional machine learning models such as XGBoost.

The inclusion of a binary classification task for prolonged hospital stays was inspired by Med-BERT (2021)^27^ suggestion to substitute the generic Next Sentence Prediction (NSP) task with more contextually relevant tasks for EHR data. Moreover, in our approach, we adopted the strategy of including a [CLS] token––following the work of Li et al. (2020)^21^ ––at the beginning of each input sequence, leveraging its role as a summarizing representation of the sequence. The [CLS] token’s aggregated representation provides a distilled feature vector that encapsulates the contextual information from the entire sequence, which is pivotal for downstream classification tasks. Additionally, the differing embedding layer designs in earlier BERT models for EHR data emphasize the necessity to investigate how these designs affect the model’s ability to accurately represent EHR temporality. By conducting ablation experiments, we evaluated the impact of integrating temporal factors—patient age, visit sequence, patient age at encounter, and inter-visit durations —on the predictive accuracy for diabetic complications.

We combined AUROC scores with Shannon’s information entropy to evaluate model performance, optimizing a balance between model simplicity and performance accuracy. We found that simpler temporal embeddings, such as visit number and inter-visit durations, offered an optimal trade-off, achieving high predictive accuracy, which is likely attributable to the information gain rooted in its representation. Combining input with visit number and inter-visit durations achieved the highest micro-averaged AUROC of 0.713 in predicting the four outcomes. This represents an improvement of 4.8% and 3.3% over the respective finetuning frameworks and a substantial 27.0% over the AUROC achieved by XGBoost. Notably, within Top-BERT, this embedding combination surpassed the AUROCs of BEHRT’s and Med-BERT’s distinct embedding designs by 5.22% and 0.28%, respectively. Top-BERT also showed improved performance over the finetuning frameworks of BEHRT and Med-BERT (enhancing AUCs by margins of 9.6% & 6.6% and 5.83% & 0.55%, respectively). Furthermore, Top-BERT showed consistent capability in handling the class imbalance with higher predictive true positive rates than XGBoost-further underscoring the potential of Top-BERT in discriminating between clinical tasks, especially in limited sample cases.

The combined use of self-supervised and supervised learning methods within Top-BERT utilizes the distinct benefits of multitask approach. Unsupervised learning methods identify underlying patterns in large volumes of unlabeled data, while supervised learning refines this understanding by directing the model’s focus toward accurately predicting designated clinical outcomes. This approach gives Top-BERT the unique ability to derive generalized insights from data and perform clinical task predictions independent of any pre-established knowledge base from pretrained models. Multitask learning has been shown to obtain a more robust shared representation of the tasks that effectively can mitigate the sparsity of labeled data, enhancing model performance, faster model convergence, and reducing overfitting risks^46^. This accounts for the case that Top-BERT outperformed the two finetuning frameworks without relying on class balancing techniques to address label sparsity in the dataset, whereas the finetuning frameworks required weighted batch samplers to improve their learning. Moreover, Top-BERT’s unified multitask learning approach achieves time efficiency and optimized performance, reaching convergence within 350 epochs (20-24 hours of training), compared to the pretraining-finetuning framework to our single-center EHR data which required approximately 28-30 hours for pretraining (epochs = 500) and an additional 8-10 hours for fine-tuning. This increase in efficiency, especially with limited data, further underscores the effectiveness of our approach.

### Common Predictive Risk Factors and Clinical Relevance

The insights from our model explanations reveal the significant predictive diagnoses used by our model to make predictions for each diabetes-related complications, including chronic kidney disease, major adverse cardiac events (MACE), retinopathy, and neuropathy. Hypertension, prevalent in approximately 69% of our cohort, was found to be a key predictive factor for all studied outcomes across all age groups (Figures 6 and 7). Our findings resonate with established clinical evidence linking hypertension to an escalated risk of diabetic complications^47,48^. Furthermore, hypertension with diabetes is associated with a 6-fold increase in the risk of cardiovascular events, a risk that escalates further with the coexistence of chronic kidney diseases^48^. Additionally, our study identified hyperlipidemia and obesity as contributory to organ damage––reinforcing their role in the pathogenesis of diabetes-related complications^49^.

Metabolic imbalances such electrolyte balance disorders—specifically potassium, magnesium, and phosphate—emerged as significant predictors across all complications, reflective of the complex interplay in diabetes management (Figure 6)^50,51^. Diabetes-related electrolyte imbalances stem from renal issues, absorption problems, acid-base imbalances, and extensive medication use. Low serum magnesium is linked to key diabetes complications, including retinopathy and heart disease, while polypharmacy can lead to hypopotassemia, further increasing cardiovascular risk. Our findings also showed association of vitamin D with both micro and macro-vascular complications, as well as identified anemia a significant predictive factor, corroborating existing clinical evidence^51, 52^.

The age-specific patterns of symptoms suggest that for younger individuals, addressing modifiable risk factors such as hyperlipidemia and obesity could be crucial in reducing the risk of diabetes-related complications. Tailoring interventions for middle-aged individuals by monitoring glucose and addressing lifestyle factors like alcohol consumption while managing lipid levels in older adults may be vital. Thus, the common risk factors identified in this study highlight the significance of regular monitoring of glucose level, blood pressure, serum electrolytes and vitamin levels, hemoglobin, lipid profile, and weight in diabetes patients to improve their overall healthcare outcomes and hence prevent the onset of other complications^53,54^.

### Symptoms of Complications and Clinical Implications

Our findings delineate a trajectory of early clinical manifestations detectable in EHR data associated with an increasing risk of the studied complications. For example, patients at elevated risk for chronic kidney disease had earlier encounters indicating potential kidney dysfunction, including symptoms like limb swelling, increased frequency of urination, and fluctuating blood pressure. Patients at increased risk for cardiovascular complications presented with symptoms such as abnormal electrocardiogram readings and various cardiac anomalies, while those at risk for neuropathy exhibited chronic pain alongside other neurological symptoms. Similarly, individuals facing a risk of retinopathy had historical clinical encounters related to eye conditions. These insights highlight the essential role of proactive symptom assessment and the management of comorbid conditions in preventing the advancement of complications in patients with diabetes, emphasizing the medical necessity for comprehensive evaluation as a fundamental element of preventive care strategies^54^.

### Limitations and Future Goals

Although our study achieved considerable progress in predicting clinical outcomes from EHR data enhancing BERT framework, we recognize several limitations that open opportunities for future research.

#### Integration of Numerical Features

Our future goal is to improve the model performance in predicting the diabetic complications, especially leading to higher AUC values. Prior studies have shown to improve model performance with inclusion of numerical features such labs and vitals (body mass index, blood pressure, lipid profiles, etc.)^15,55,56^. BERT inherently processes inputs as tokens, including numerical features. This presents a limitation as it does not fully exploit the quantitative nature of these features. Thus, we will focus on integrating numerical data, such as lab values and vital signs, more effectively within our model architecture, which may potentially lead to enhanced model performance and identification of modifiable predictive factors.

#### Model Fairness Studies

Our research also highlights the role of thorough documentation and patterns of healthcare utilization in the EHR, which significantly contribute to the predictive features of our models. For example, patients with higher healthcare utilization tend to accumulate more diagnostic entries in their EHRs, influencing the model’s predictions. As AI and machine learning become more prevalent in healthcare, it’s critical to maintain model integrity and prevent biases that could adversely affect certain groups of patients. In our future work, we aim to conduct thorough model fairness studies to identify and mitigate any biases and confounding factors, ensuring equitable predictions across all demographics.

#### Multi-center Data Expansion

The current study’s findings are based on single-center data, which may not capture the diversity of patient populations and practice patterns. Expanding the dataset to include multiple centers will allow our model to learn from a broader range of patient encounters, enhancing its generalizability and robustness across different clinical environments.

Thus, addressing these limitations is pivotal for advancing our research. The envisioned improvements and expansions will aim not just to refine the predictive accuracy but also to ensure that the insights generated by our model are equitable, generalizable, and applicable across various clinical settings.

## Conclusion

In summary, Top-BERT introduces a transformative approach that deviates from the traditional pretraining-finetuning paradigm of language models, demonstrating its effective predictive performance on clinical tasks within the confines of a single-center EHR dataset, even with limited sample sizes. The model explanations using Integrated Gradients further validate the clinical applicability of our findings, highlighting its promise for enhancing diabetes care management and patient health outcomes. Moreover, our study deployed a robust framework integrating the strengths of MU EHR data lake and NRP Nautilus infrastructures––leveraging parallel high-performance computing significantly reduced model training time, boosting efficiency, and accelerating the development of our sophisticated predictive models from an estimated 65 days to just 2 days. Furthermore, we prioritized the reproducibility of our research, making our codes accessible and clearly illustrating our methodology through informative graphics and diagrams.

## Data Availability

All data produced are obtained from the University of Missouri PCORnet Common Data Model.

## Code Availability

The complete codebase for this framework is accessible through our GitHub repository: https://github.com/hikf3/task-oriented-predictive-BERT

## Acknowledgements

The data set used for the analyses described were obtained from PCORnet Common Data Model at the University of Missouri, which is funded by the Patient Centered Outcomes Research Institute PCORnet (RI-MISSOURI-01-PS1) Clincal Research Network, the Greater Plains Collaborative. The NSF Nautilus Kubernetes HyperCluster is supported by the University of Missouri Greater Plains Regional CyberTeam (NSF Award #1925681).

## Author contributions

**HI:** conceptualization, methodology, data analysis, clinical interpretations, and writing.

**GB:** clinical interpretations, validation, review, and editing.

**RP:** clinical interpretations, validation, review, and editing.

**PR:** methodology, validation, review, and editing.

**RW:** clinical interpretations, validation, review, and editing.

**XS:** supervision, methodology, clinical interpretations, validation, review, and editing.

## Supplementary Document

### Comparing BERT-based Model Performances

Table S1 shows the micro-averaged performance metrics for two finetuning frameworks and Top-BERT embedding configurations. Metrics were computed for epoch 200 for the two finetuning frameworks after pretraining for 500 epochs. For the Top-BERT models all metrics were computed at epoch 350. We also compared the AUROC of prolonged hospital stay in the Top-BERT models, as shown in Table S1.

**Table S 1.**
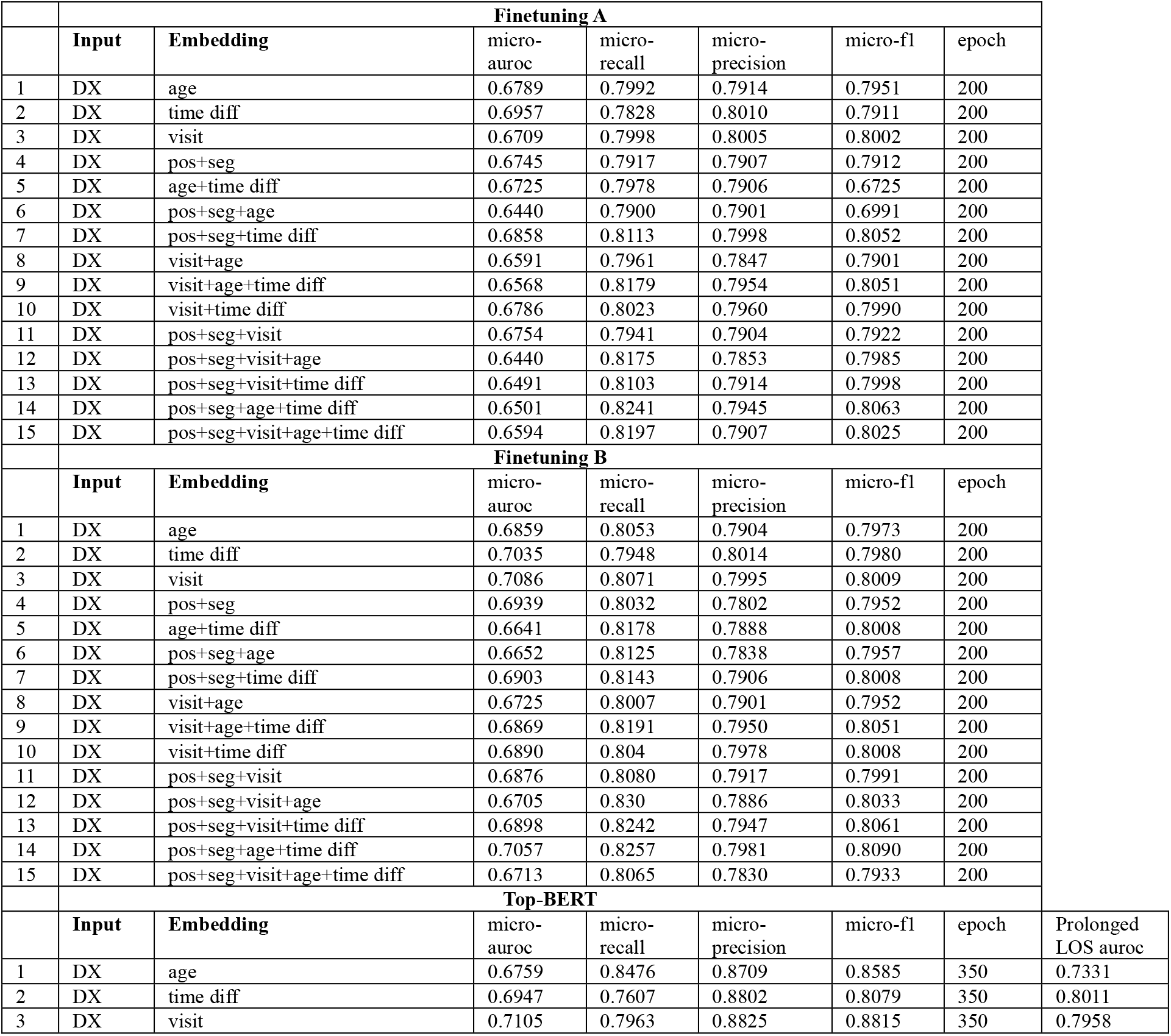
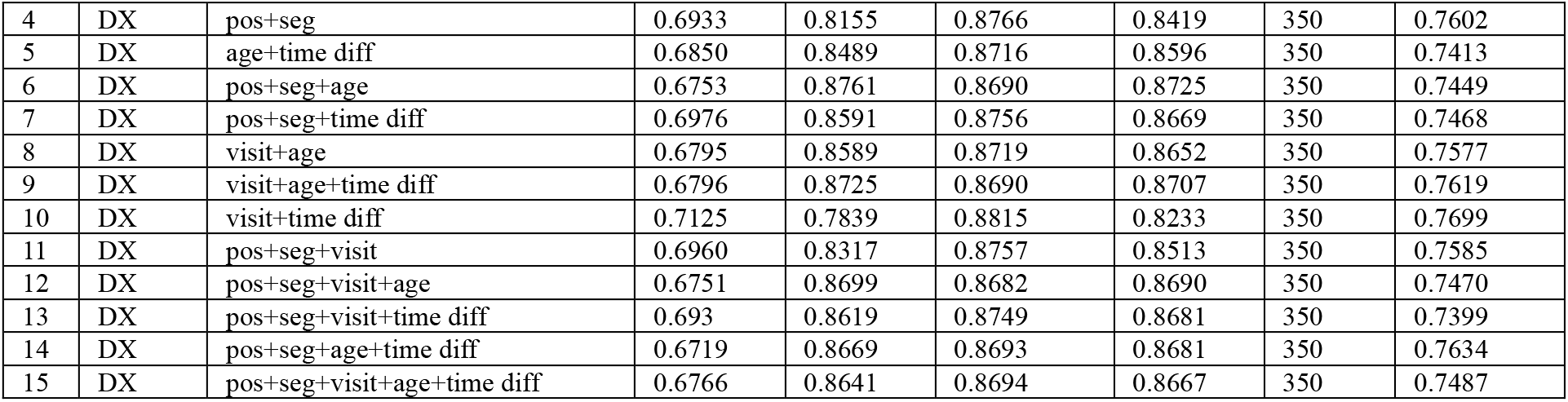
Comparing the micro-averaged metrics for each embedding configurations for the BERT-based model frameworks. Additionally, the AUROC of prolonged hospital stay is provided for the Top-BERT models.

### Comparing Task-Specific Model Performances

To gain insights on the differences and similarities of Top-BERT and XGBoost in handling class imbalance, we compared the model performance for each classification task: CKD, MACE, NEUR, and RET using precision, recall, F1-score, Mathew’s correlation coefficient (MCC) and confusion matrix computed at varying threshold values (10%, 30%, 50%, and 80%). Our test data consisted of 7.25% of CKD labels, 11.6% of MACE, 10.9% of NEUR, and 2.7% of RET. Table S2 compares the metrics for each classification task for five Top-BERT embedding configurations (having comparable mAUCs) and XGBoost. Top-BERT models displayed more balanced metrics across the thresholds with significantly higher true positive predictions, leading to higher recall values than XGBoost. However, XGBoost showed lower false positive rates, contributing to higher precision, particularly at higher thresholds than Top-BERT. For instance, XGBoost achieved a precision of 69.5% for predicting MACE at 50% threshold compared to 26.4% in Top-BERT (input+time_diff). False positives of MACE predicted by XGBoost were significantly lower than Top-BERT models (0.28% for XGBoost vs. 9.32% for Top-BERT input+time_diff) –– which contributed to the significant increase in precision for XGBoost. However, XGBoost identified 41 true positives out of the 849 positive labels, which resulted in a recall value of 4.8% compared to 414 true positives (recall of 48.8%) for Top-BERT (input+time_diff). Additionally, to predict RET, which had the lowest fraction of true positives, true positive rates for XGBoost were zero in most cases, whereas input+time_diff identified 21.5% true positives at the 50% threshold. Thus, we observed that higher precision values and lower recall values obtained by XGBoost were consequences of conservatively predicting fewer number of positive cases (both true positives and false positives) in all classification tasks. On the other hand, Top-BERT showed better measure of separability, as indicated by higher mAUC values, and higher efficiency in predicting the positive classes compared to XGBoost.

**Table S 2.**
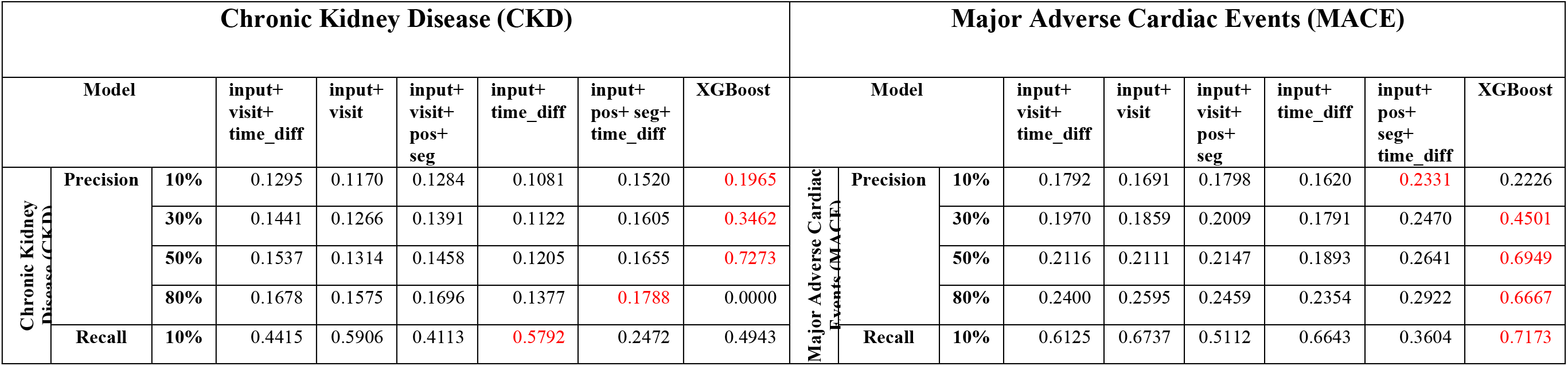
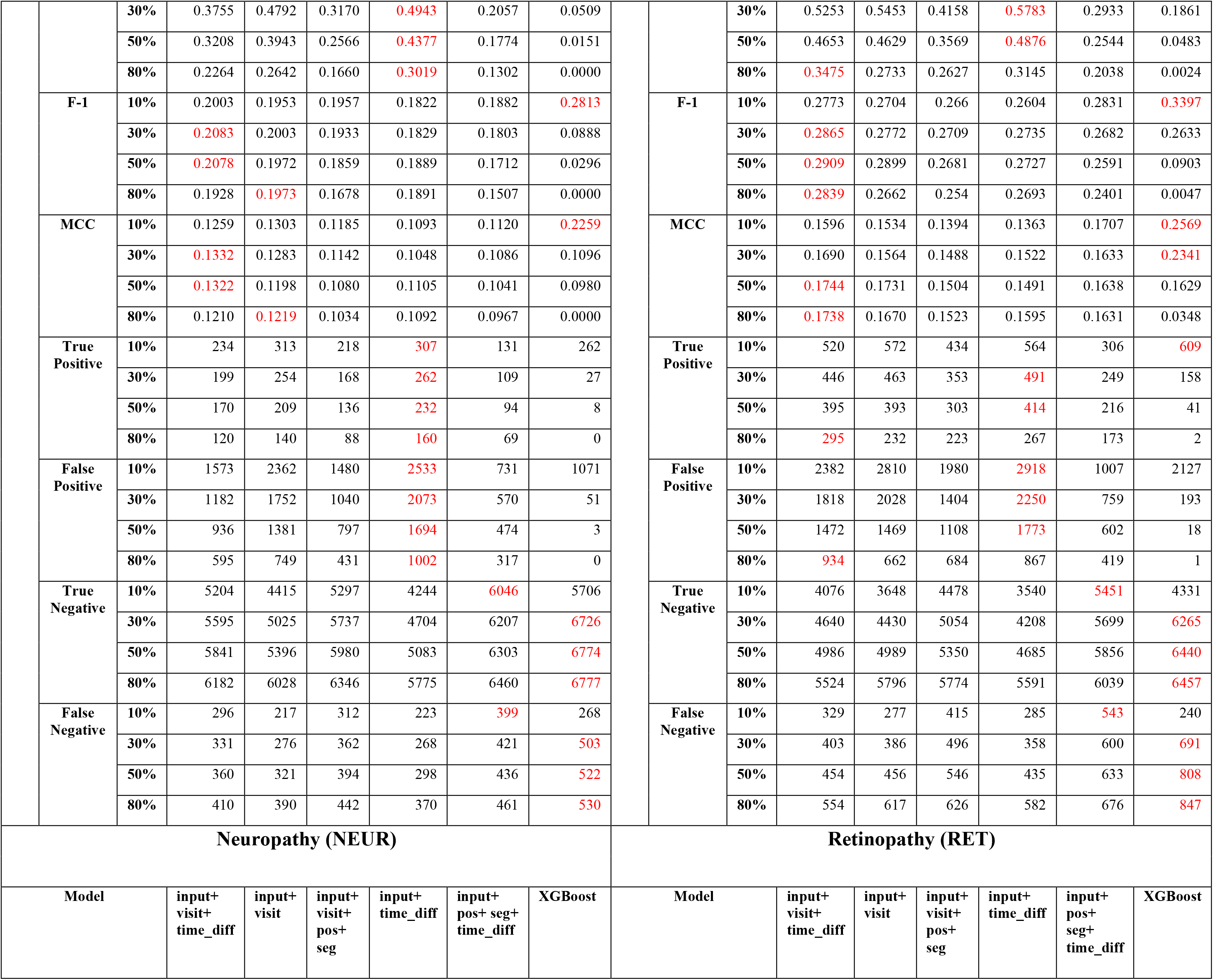
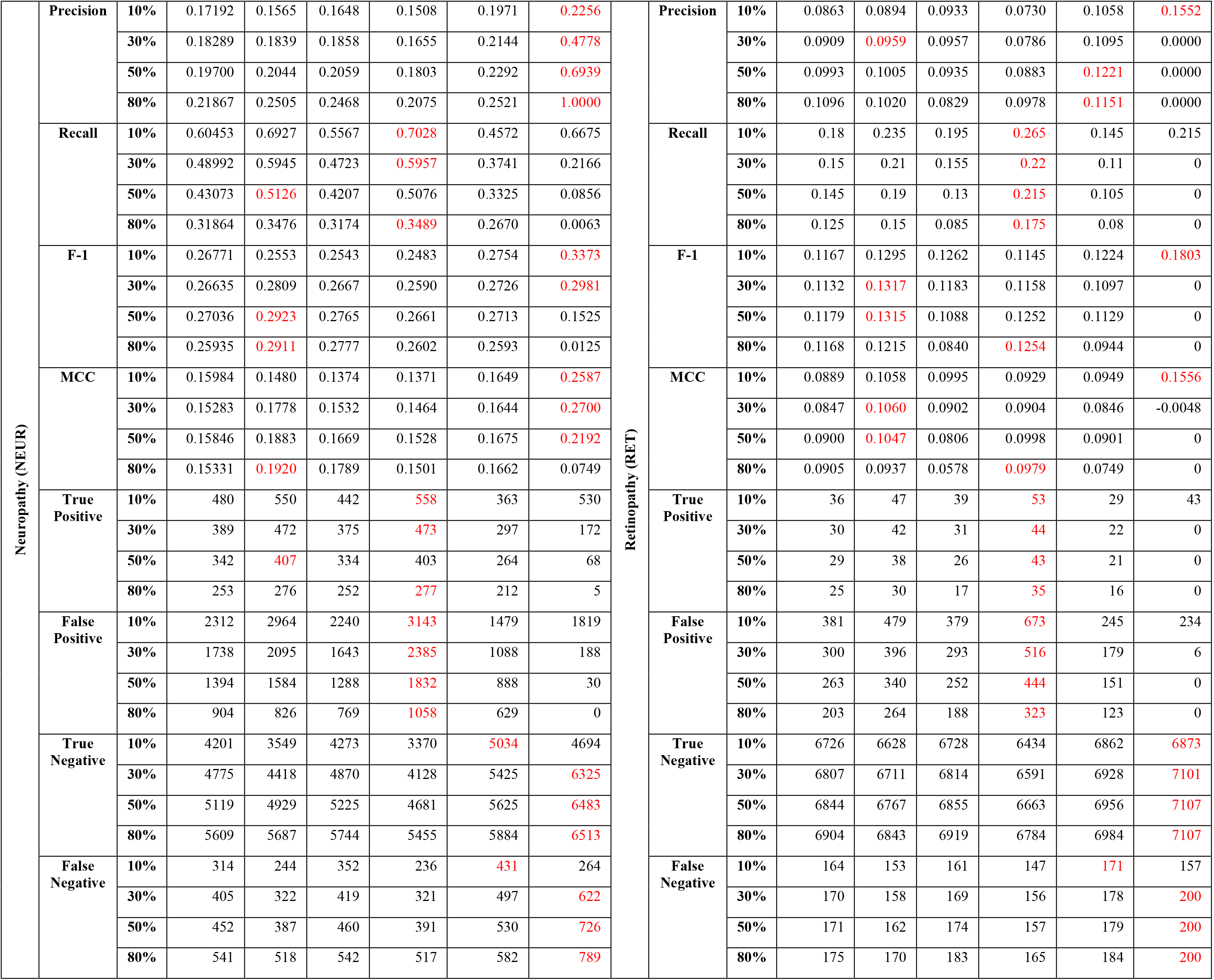
Comparing the task-specific model performances for five Top-BERT embedding configurations (having comparable micro-averaged AUROCs) and XGBoost for each classification task: CKD, MACE, NEUR, and RET using precision, recall, F1-score, Mathew’s correlation coefficient (MCC) and confusion matrix computed at varying threshold values (10%, 30%, 50%, and 80%). The red highlighted scores show the highest value in each metric for each threshold value.

